# Associations between inhibitory control, stress, and alcohol (mis)use during the first wave of the COVID-19 pandemic in the UK: a national cross-sectional study utilising data from four birth cohorts

**DOI:** 10.1101/2020.09.24.20197293

**Authors:** James M. Clay, Lorenzo D. Stafford, Matthew O. Parker

## Abstract

**Aims:** To investigate: (1) alcohol use during the pandemic in the UK; and (2) the extent to which poor inhibitory control and/or stress were associated with any change in alcohol use or hazardous drinking.

**Design:** Cross-sectional online survey administered between 2 and 31 May 2020.

**Setting:** UK.

**Participants:** 13,453 respondents aged 19 – 62 years comprising participants of four nationally representative birth cohorts (19, 30, 50 and 62-years old).

**Measurements:** Change in alcohol use and risk of alcohol-related harm due to hazardous drinking and change in stress since the start of the Coronavirus outbreak; inhibitory control (impatience and risk-taking); and sociodemographic characteristics, including diagnosed or suspected COVID-19, and key worker status.

**Findings:** Most respondents reported consuming/feeling the same amount or less alcohol/stress. However, a significant minority, particularly among thirty-(29.08%) and fifty-year-olds (26.67%), reported drinking more, and between 32.23% and 45.02% of respondents reported feeling more stressed depending on cohort. Being female was associated with an increased likelihood of reporting heightened stress (OR_19_ = 1.54, 95% CI 1.08 to 2.20; OR_30_ = 1.93, 95% CI 1.39 to 2.70; OR_50_ = 1.62, 95% CI 1.37 to 1.92; OR_62_ = 2.03, 95% CI 1.66 to 2.48). Stress was associated with hazardous drinking among 30-year-olds (OR = 3.77, 95% CI 1.15 to 12.28). Impatience was associated with both increased alcohol use (1.14, 95% CI 1.06, 1.24) and hazardous drinking (1.20, 95% CI 1.05, 1.38) among 19-year-olds. Risk-taking was associated with hazardous drinking for 30-year-olds (OR = 1.18, 95% CI 1.05, 1.32).

**Conclusions:** These data highlight concerns about the UK government’s stance on the ‘essential’ nature of off-premises alcohol sales during the lockdown, particularly from a public health perspective, when considering those at risk of alcohol misuse and alcohol-related harm.

## Introduction

Since being first identified in Wuhan, China, in December 2019, severe acute respiratory syndrome coronavirus 2 (SARS-CoV-2), the virus that causes COVID-19, has caused a significant threat to global health [1]. Governments around the world responded by imposing ‘lockdowns’ (orders to remain at home, and socially isolate) on their populations, and available evidence supports this action as a means of mitigating the rate of spread of the virus [2]. However, the indirect impact of lockdown on public health has raised concern, particularly relating to mental health and wellbeing [3].

Concerns that the lockdowns may increase alcohol misuse have been raised, particularly concerning people at high-risk of developing, or re-establishing, hazardous alcohol use [4–6]. An example of individuals who are at high risk of alcohol misuse are people that display poor inhibitory control [7,8]. Inhibitory control is generally conceptualised as one of the core executive functions [9]. It is a complex and multifaceted construct made up of several subcomponents: *<u>response inhibition</u>* (i.e., action inhibition, action cancellation), *<u>sensitivity to delay</u>* (i.e., delay discounting, patience), *<u>sensitivity to risk/reward</u>* (risk-taking, sensation seeking), and *<u>attention</u>* (i.e., capacity to focus and avoid interference) [10]. Indeed, several lines of evidence from pre-clinical translational work [11,12], neuroimaging studies [13,14], and heritability studies [15,16] converge to suggest that poor inhibitory control is both a risk factor for the development, and consequence, of substance misuse and addiction.

The association between stress and alcohol use is also well established [17–19]. Similar to inhibitory control, stress plays a critical role in both the onset and maintenance of alcohol misuse and addiction [20]. On the one hand, the acute anxiolytic properties of alcohol motivate some individuals to drink [21]. On the other, perhaps counterintuitively, alcohol acts as a physiological ‘stressor’: acute exposure to alcohol stimulates the hypothalamic-pituitary-adrenocortical (HPA) axis through direct activation of GABA_A_ receptors in the paraventricular nucleus [22]. Finally, exposure to either chronic stress or chronic alcohol misuse both lead to blunted stress responses, including dysregulation of the HPA axis – a known risk factor for hazardous drinking and addiction [23].

Recently we have demonstrated a complex interplay between inhibitory control, stress, and alcohol use, where an experimentally induced acute psychosocial stressor increased craving for alcohol [24], and voluntary alcohol consumption[25] in healthy (non-addicted) individuals. We found that the strength of these stress–induced increases in alcohol craving and consumption were predicated on individual differences in risk-taking personality traits, stress-reactivity, and stress-recovery. Collectively, our findings suggest these innate (e.g., poor inhibitory control), and environmental (e.g., ‘state’ induced stress) factors may combine to make particular individuals more at risk of alcohol misuse.

Here, we analysed the first sweep of the Centre for Longitudinal Studies (CLS) COVID-19 survey [26] – which was answered by individuals from five nationally representative cohorts who have been providing data since childhood – to investigate: (1) alcohol use during the pandemic in the UK; and (2) the extent to which poor inhibitory control and/or stress were associated with any change in alcohol use or hazardous drinking.

## Methods

### Data source

We used data from the first wave of the CLS COVID-19 survey [26]. The survey design, recruitment procedure, and fieldwork processes have been described in detail elsewhere [27]. Briefly, the survey was administered between 2 and 31 May 2020, using Qualtrics (Provo, Utah), to 50,479 individuals from five nationally representative UK birth cohorts. These included: (1) the Millennium Cohort Study (MCS), who are part of ‘Generation Z’, and were aged 19; (2) Next Steps, who are part of the ‘Millennial’ generation, who were aged 30; (3) the 1970 British Cohort Study (BCS70), who belong to ‘Generation X’ – aged 50; (4) the National Child Development Study (NCDS), who were aged 62 and were born in the latter part of the ‘Baby Boomer’ generation; and (5) the National Study of Health and Development (NSHD), who were born at the beginning of the ‘Baby Boomer’ era, and were aged Due to the nature of the survey, only those who had their email address previously recorded were approached. Overall, 18,042 of those invited responded, achieving a response rate (RR) of 35.7%. This response rate is similar to comparable web surveys conducted at this time, such as the Understanding Society COVID-19 survey [28]. Ethnicity data was linked from previous survey waves [29–32]. All data used in this study are available from the UK Data Service Website (https://ukdataservice.ac.uk/) under the “Safeguarded” data access policy. Approval for the present study was obtained from the University of Portsmouth. Informed consent from participants was sought by the CLS team.

### Study sample

Due to data availability at the time of analysis, four of the five cohorts included in the COVID-19 survey were analysed. Namely, the MCS cohort members (*n* = 2,645, RR = 26.59%), Next Steps (*n* = 1,907, RR = 20.33%), the BCS70 (*n* = 4,223, RR = 40.38%), and the NCDS (*n* = 5,178, RR = 57.90%). The study was restricted to UK-based respondents; thus emigrants (*n* = 500) were excluded prior to analysis. This left 13,453 cases for analysis. A detailed overview of the study sample is presented in the Supporting Information, Figure S1. Selected sample characteristics are shown in Table 1.

**Table 1.**
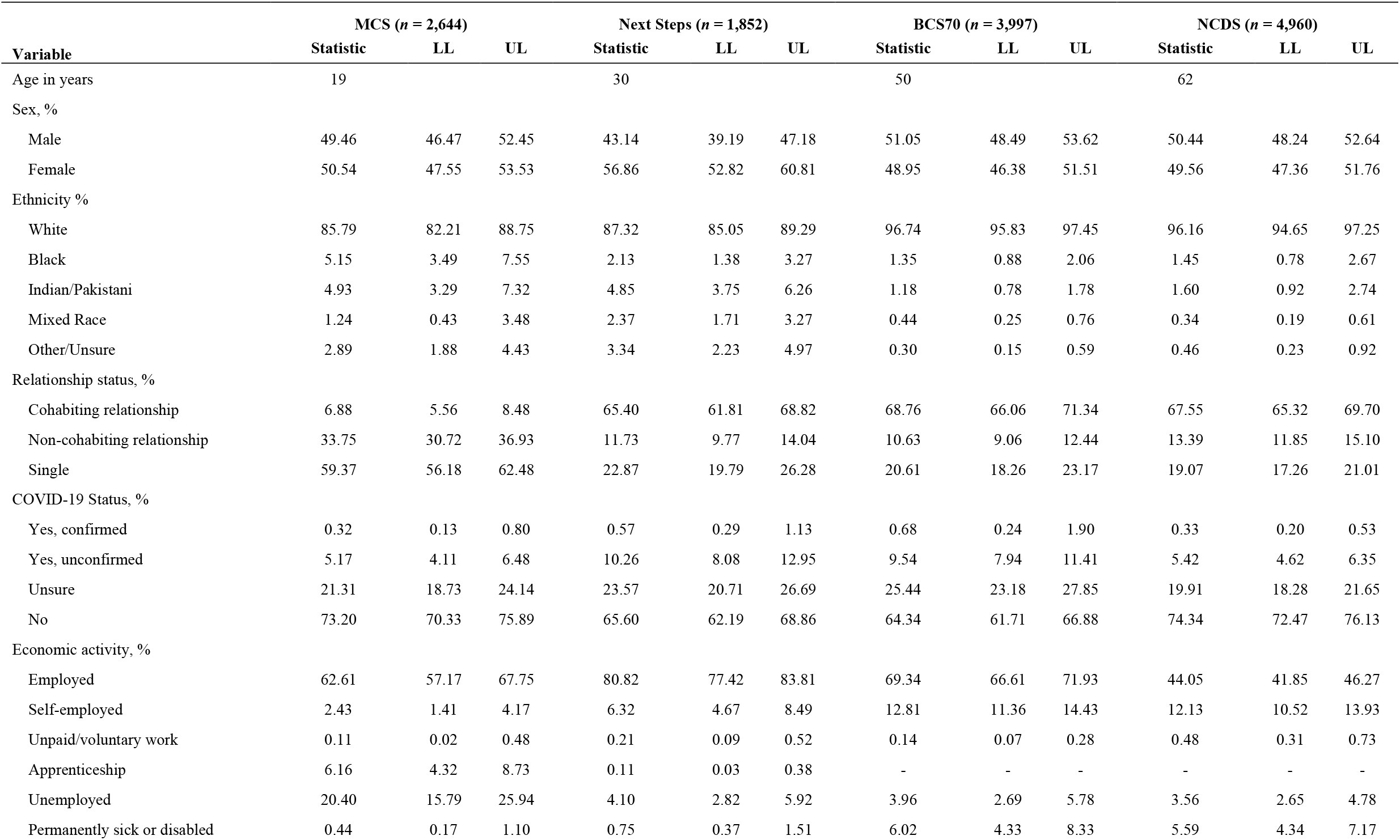
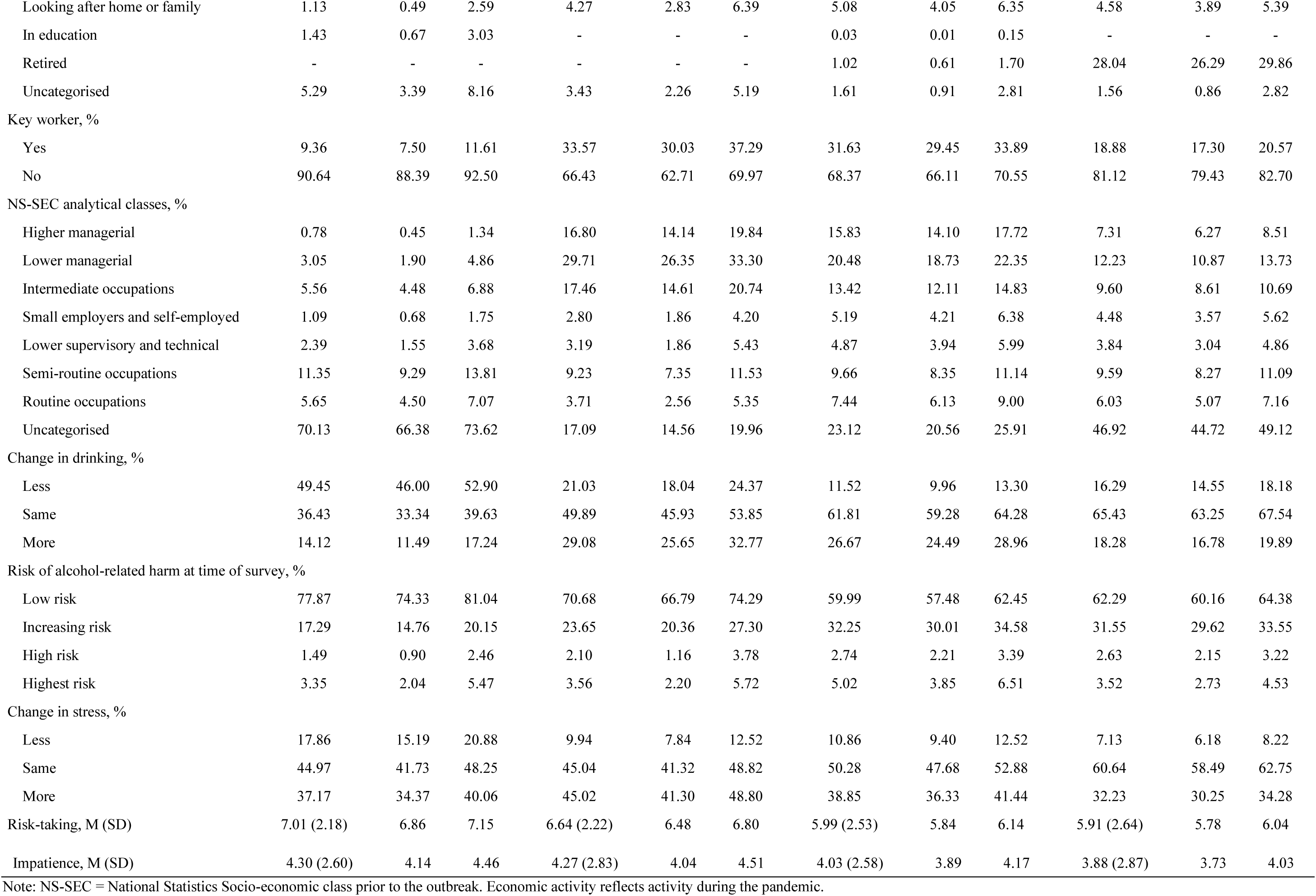
Selected demographic characteristics.

| Variable | MCS (*n* = 2,644) |  |  | Next Steps (*n* = 1,852) |  |  | BCS70 (*n* = 3,997) |  |  | NCDS (*n* = 4,960) |  |  |
| --- | --- | --- | --- | --- | --- | --- | --- | --- | --- | --- | --- | --- |
|  | Statistic | LL | UL | Statistic | LL | UL | Statistic | LL | UL | Statistic | LL | UL |
| Age in years | 19 |  |  | 30 |  |  | 50 |  |  | 62 |  |  |
| Sex, % |  |  |  |  |  |  |  |  |  |  |  |  |
| Male | 49.46 | 46.47 | 52.45 | 43.14 | 39.19 | 47.18 | 51.05 | 48.49 | 53.62 | 50.44 | 48.24 | 52.64 |
| Female | 50.54 | 47.55 | 53.53 | 56.86 | 52.82 | 60.81 | 48.95 | 46.38 | 51.51 | 49.56 | 47.36 | 51.76 |
| Ethnicity % |  |  |  |  |  |  |  |  |  |  |  |  |
| White | 85.79 | 82.21 | 88.75 | 87.32 | 85.05 | 89.29 | 96.74 | 95.83 | 97.45 | 96.16 | 94.65 | 97.25 |
| Black | 5.15 | 3.49 | 7.55 | 2.13 | 1.38 | 3.27 | 1.35 | 0.88 | 2.06 | 1.45 | 0.78 | 2.67 |
| Indian/Pakistani | 4.93 | 3.29 | 7.32 | 4.85 | 3.75 | 6.26 | 1.18 | 0.78 | 1.78 | 1.60 | 0.92 | 2.74 |
| Mixed Race | 1.24 | 0.43 | 3.48 | 2.37 | 1.71 | 3.27 | 0.44 | 0.25 | 0.76 | 0.34 | 0.19 | 0.61 |
| Other/Unsure | 2.89 | 1.88 | 4.43 | 3.34 | 2.23 | 4.97 | 0.30 | 0.15 | 0.59 | 0.46 | 0.23 | 0.92 |
| Relationship status, % |  |  |  |  |  |  |  |  |  |  |  |  |
| Cohabiting relationship | 6.88 | 5.56 | 8.48 | 65.40 | 61.81 | 68.82 | 68.76 | 66.06 | 71.34 | 67.55 | 65.32 | 69.70 |
| Non-cohabiting relationship | 33.75 | 30.72 | 36.93 | 11.73 | 9.77 | 14.04 | 10.63 | 9.06 | 12.44 | 13.39 | 11.85 | 15.10 |
| Single | 59.37 | 56.18 | 62.48 | 22.87 | 19.79 | 26.28 | 20.61 | 18.26 | 23.17 | 19.07 | 17.26 | 21.01 |
| COVID-19 Status, % |  |  |  |  |  |  |  |  |  |  |  |  |
| Yes, confirmed | 0.32 | 0.13 | 0.80 | 0.57 | 0.29 | 1.13 | 0.68 | 0.24 | 1.90 | 0.33 | 0.20 | 0.53 |
| Yes, unconfirmed | 5.17 | 4.11 | 6.48 | 10.26 | 8.08 | 12.95 | 9.54 | 7.94 | 11.41 | 5.42 | 4.62 | 6.35 |
| Unsure | 21.31 | 18.73 | 24.14 | 23.57 | 20.71 | 26.69 | 25.44 | 23.18 | 27.85 | 19.91 | 18.28 | 21.65 |
| No | 73.20 | 70.33 | 75.89 | 65.60 | 62.19 | 68.86 | 64.34 | 61.71 | 66.88 | 74.34 | 72.47 | 76.13 |
| Economic activity, % |  |  |  |  |  |  |  |  |  |  |  |  |
| Employed | 62.61 | 57.17 | 67.75 | 80.82 | 77.42 | 83.81 | 69.34 | 66.61 | 71.93 | 44.05 | 41.85 | 46.27 |
| Self-employed | 2.43 | 1.41 | 4.17 | 6.32 | 4.67 | 8.49 | 12.81 | 11.36 | 14.43 | 12.13 | 10.52 | 13.93 |
| Unpaid/voluntary work | 0.11 | 0.02 | 0.48 | 0.21 | 0.09 | 0.52 | 0.14 | 0.07 | 0.28 | 0.48 | 0.31 | 0.73 |
| Apprenticeship | 6.16 | 4.32 | 8.73 | 0.11 | 0.03 | 0.38 | - | - | - | - | - | - |
| Unemployed | 20.40 | 15.79 | 25.94 | 4.10 | 2.82 | 5.92 | 3.96 | 2.69 | 5.78 | 3.56 | 2.65 | 4.78 |
| Permanently sick or disabled | 0.44 | 0.17 | 1.10 | 0.75 | 0.37 | 1.51 | 6.02 | 4.33 | 8.33 | 5.59 | 4.34 | 7.17 |
| Looking after home or family | 1.13 | 0.49 | 2.59 | 4.27 | 2.83 | 6.39 | 5.08 | 4.05 | 6.35 | 4.58 | 3.89 | 5.39 |
| In education | 1.43 | 0.67 | 3.03 | - | - | - | 0.03 | 0.01 | 0.15 | - | - | - |
| Retired | - | - | - | - | - | - | 1.02 | 0.61 | 1.70 | 28.04 | 26.29 | 29.86 |
| Uncategorised | 5.29 | 3.39 | 8.16 | 3.43 | 2.26 | 5.19 | 1.61 | 0.91 | 2.81 | 1.56 | 0.86 | 2.82 |
| Key worker, % |  |  |  |  |  |  |  |  |  |  |  |  |
| Yes | 9.36 | 7.50 | 11.61 | 33.57 | 30.03 | 37.29 | 31.63 | 29.45 | 33.89 | 18.88 | 17.30 | 20.57 |
| No | 90.64 | 88.39 | 92.50 | 66.43 | 62.71 | 69.97 | 68.37 | 66.11 | 70.55 | 81.12 | 79.43 | 82.70 |
| NS-SEC analytical classes, % |  |  |  |  |  |  |  |  |  |  |  |  |
| Higher managerial | 0.78 | 0.45 | 1.34 | 16.80 | 14.14 | 19.84 | 15.83 | 14.10 | 17.72 | 7.31 | 6.27 | 8.51 |
| Lower managerial | 3.05 | 1.90 | 4.86 | 29.71 | 26.35 | 33.30 | 20.48 | 18.73 | 22.35 | 12.23 | 10.87 | 13.73 |
| Intermediate occupations | 5.56 | 4.48 | 6.88 | 17.46 | 14.61 | 20.74 | 13.42 | 12.11 | 14.83 | 9.60 | 8.61 | 10.69 |
| Small employers and self-employed | 1.09 | 0.68 | 1.75 | 2.80 | 1.86 | 4.20 | 5.19 | 4.21 | 6.38 | 4.48 | 3.57 | 5.62 |
| Lower supervisory and technical | 2.39 | 1.55 | 3.68 | 3.19 | 1.86 | 5.43 | 4.87 | 3.94 | 5.99 | 3.84 | 3.04 | 4.86 |
| Semi-routine occupations | 11.35 | 9.29 | 13.81 | 9.23 | 7.35 | 11.53 | 9.66 | 8.35 | 11.14 | 9.59 | 8.27 | 11.09 |
| Routine occupations | 5.65 | 4.50 | 7.07 | 3.71 | 2.56 | 5.35 | 7.44 | 6.13 | 9.00 | 6.03 | 5.07 | 7.16 |
| Uncategorised | 70.13 | 66.38 | 73.62 | 17.09 | 14.56 | 19.96 | 23.12 | 20.56 | 25.91 | 46.92 | 44.72 | 49.12 |
| Change in drinking, % |  |  |  |  |  |  |  |  |  |  |  |  |
| Less | 49.45 | 46.00 | 52.90 | 21.03 | 18.04 | 24.37 | 11.52 | 9.96 | 13.30 | 16.29 | 14.55 | 18.18 |
| Same | 36.43 | 33.34 | 39.63 | 49.89 | 45.93 | 53.85 | 61.81 | 59.28 | 64.28 | 65.43 | 63.25 | 67.54 |
| More | 14.12 | 11.49 | 17.24 | 29.08 | 25.65 | 32.77 | 26.67 | 24.49 | 28.96 | 18.28 | 16.78 | 19.89 |
| Risk of alcohol-related harm at time of survey, % |  |  |  |  |  |  |  |  |  |  |  |  |
| Low risk | 77.87 | 74.33 | 81.04 | 70.68 | 66.79 | 74.29 | 59.99 | 57.48 | 62.45 | 62.29 | 60.16 | 64.38 |
| Increasing risk | 17.29 | 14.76 | 20.15 | 23.65 | 20.36 | 27.30 | 32.25 | 30.01 | 34.58 | 31.55 | 29.62 | 33.55 |
| High risk | 1.49 | 0.90 | 2.46 | 2.10 | 1.16 | 3.78 | 2.74 | 2.21 | 3.39 | 2.63 | 2.15 | 3.22 |
| Highest risk | 3.35 | 2.04 | 5.47 | 3.56 | 2.20 | 5.72 | 5.02 | 3.85 | 6.51 | 3.52 | 2.73 | 4.53 |
| Change in stress, % |  |  |  |  |  |  |  |  |  |  |  |  |
| Less | 17.86 | 15.19 | 20.88 | 9.94 | 7.84 | 12.52 | 10.86 | 9.40 | 12.52 | 7.13 | 6.18 | 8.22 |
| Same | 44.97 | 41.73 | 48.25 | 45.04 | 41.32 | 48.82 | 50.28 | 47.68 | 52.88 | 60.64 | 58.49 | 62.75 |
| More | 37.17 | 34.37 | 40.06 | 45.02 | 41.30 | 48.80 | 38.85 | 36.33 | 41.44 | 32.23 | 30.25 | 34.28 |
| Risk-taking, M (SD) | 7.01 (2.18) | 6.86 | 7.15 | 6.64 (2.22) | 6.48 | 6.80 | 5.99 (2.53) | 5.84 | 6.14 | 5.91 (2.64) | 5.78 | 6.04 |
| Impatience, M (SD) | 4.30 (2.60) | 4.14 | 4.46 | 4.27 (2.83) | 4.04 | 4.51 | 4.03 (2.58) | 3.89 | 4.17 | 3.88 (2.87) | 3.73 | 4.03 |

Note: NS-SEC = National Statistics Socio-economic class prior to the outbreak. Economic activity reflects activity during the pandemic.

### Outcome measures

Alcohol use behaviour was assessed using five questions from the Alcohol Use Disorders Identification Test (AUDIT) -a tool developed by the World Health Organisation as a brief assessment of alcohol misuse [33]. The original AUDIT has been shown to have excellent psychometric properties when used to assess alcohol use disorders in a variety of settings including both college students [34] and during routine health examinations [35].

The questions administered during the survey were:

1. “How often have you had a drink containing alcohol?”
2. “How many standard alcoholic drinks have you had on a typical day when you were drinking?”
3. “How often have you found you were not able to stop drinking once you had started?”
4. “How often have you failed to do what was expected of you because of drinking?”
5. “Has a relative, friend, doctor, or health worker been concerned about your drinking or advised you to cut down?”

Questions one and two were repeated, prefaced by either “in the month before the Coronavirus outbreak”, or “since the start of the Coronavirus outbreak”. This provided an assessment of alcohol use prior to, and during, the pandemic. Questions one to five were posed in the context of the pandemic, thus were worded using the latter phrasing, offering an assessment of hazardous drinking during the outbreak.

Each item was scored in line with the original AUDIT. Scores which represented alcohol use prior to and during the pandemic were calculated by summing questions one and two. A change score was calculated by subtracting the pre-pandemic from intra-pandemic score. Thus, values equal to zero reflected no change, values greater than zero represented an increase, and values less than zero denoted a reduction in alcohol use. A score representing risk of alcohol-related harm due to hazardous drinking during the pandemic was calculated by summing all items which used the latter wording. The hazardous drinking score was categorised proportionally to the original AUDIT. Whereby, a score between zero and three was coded as “Low risk”; a score between four and six was classified as “Increasing risk”; scores between seven and eight were labelled “Higher risk”; and scores of nine or greater were classed as “Highest risk”.

### Stress

Perceived stress was assessed using a single question: “Since the Coronavirus outbreak, please indicate how the following have changed… The amount of stress I’ve been feeling”. The possible responses included “More than before”, “Same – no change”, and “Less than before”. As it is well-known that experiencing symptoms of depression and/or anxiety is associated with increased psychological stress [36,37], we used linear regression models for each cohort to determine the relationship between scores on the Patient Health Questionnaire-4 (PHQ-4) [38] – an ultra-brief tool, with good psychometric properties, designed to screen for anxiety and depression in both clinical and non-clinical settings – and the stress item used here (see Table S1). After controlling for potential confounders (see below), individuals who said they were feeling more stressed than before the pandemic scored approximately two points higher (range = 1.95 – 2.68, *p*_s_ < .001) than those who said they felt the same.

### Inhibitory control

Two measures of inhibitory control were administered in the survey: patience and risk-taking. Each was measured using a single ten-point Likert scale item. The questions were phrased “On a scale from 0 – 10, where 0 is ‘never’ and 10 is ‘always’, how *willing to take risks/patient* would say you are?”. A similar single-item scale of risk preference, known as the General Risk Question (GRQ) [39], has been used extensively and has been included in several widely analysed surveys, such as the Household, Income and Labour Dynamics in Australia Survey [40], and the Understanding Society Survey [41]. Recent work suggests that the self-report (e.g., GRQ) assessment of risk-taking oftentimes outperform behavioural assessments (e.g., laboratory lotteries) due to self-report assessments taking subjective internal states, such as regret or need, into account [42]. Moreover, during the development of the Global Preferences Survey [43] – which was conducted to investigate risk and time (patience) preferences – Falk et al. [44] experimentally validated their measures by (among other things) assessing the association between single-item assessments and behavioural measures of the same constructs through Spearman’s correlations and linear regression models. Their analysis shows that the single-item assessments were moderately correlated with the behavioural measures (see Table S2). The “patience” item was reverse scored to reflect greater impatience.

### Potential confounders

Potential confounding variables were identified using the author’s substantive knowledge about established risk factors that could plausibly be related to our outcome variables. These included respondent’s sex, ethnicity, National Statistics Socio-economic Class (NS-SEC) prior to the outbreak of Coronavirus, and economic activity during the pandemic. Further information on these measures is presented in the Supporting Information.

### Statistical analysis

Statistical analysis was conducted using Stata IC (version 16.1). Figures were generated using *ggplot2* (version 3.3.2) for R (version 3.6.2) was used to create figures. Inverse probability weighting was used to account for bias introduced due to missing data, and to ensure the results were as representative as possible [45]. The overall percentage of missing data was 23.43%. The median percentage of missing data by variable was 5.29% (IQR = 8.01%). See Supporting Information and Table S3 for a detailed description of missing data. Separate analyses were conducted for each cohort due to differences in sampling methods and therefore design weights. Descriptive statistics (mean and standard deviation or proportion alongside 95% CIs) were calculated our variables of interest and select demographic variables. Prevalence estimates (with 95% CIs) split by sex, ethnicity, economic activity, and NS-SEC were calculated for our outcome measures and change in stress. Ordinal regression models were used to assess whether sub-group membership was associated with change in alcohol use, risk of alcohol-related harm due to hazardous drinking, or a change in stress levels, and to investigate associations between inhibitory control, stress, and alcohol use. We first regressed our outcome measures and change in stress on sex, ethnicity, economic activity, and NS-SEC. We then added parameters for inhibitory control, stress, and the interaction between inhibitory control and stress to our models containing our outcome variables. Given that most respondents across all cohorts were White, and since some ethnic groups made up less than one percent of the sample, a dichotomous White/non-White variable was used in regression analyses. We also noticed that the standard error among fifty-year-olds that reported being in education during the pandemic was inflated, leading to implausible results, due to only two fifty-year-olds females falling into this category. These two cases were omitted for all regression analyses, which had no impact on the final results of the models. For brevity, model estimates for potential confounders are reported in the Supporting information, Tables S4 – S15. Finally, as neither the study nor analysis plan were pre-registered on a publicly available platform, the results should be considered exploratory.

## Results

### Change in alcohol use during first lockdown

Across all cohorts, most respondents reported drinking the same amount of alcohol or less since the start of the pandemic (Table 1). Thirty-year-olds and fifty-year-olds were most likely to report increased drinking with around one-third and one-quarter reporting an increase respectively.

Figure 1 shows change in alcohol use by sub-group. In all cohorts except for sixty-two-year-olds, being employed was associated with reporting increased alcohol use (Supporting information., Tables S4 – S6). Fifty-year-old and sixty-two-year-old females had 1.27 (95% CI 1.08 to 1.50) and 1.23 (95% CI 1.02 to 1.50) times the odds of reporting increased alcohol use, respectively (Supporting information, Tables S6 and S7). Regarding socio-economic class, fifty-year-olds who worked in intermediate occupations (OR = 0.70, 95% CI 0.54 to 0.92), semi-routine occupations (OR = 0.62, 95% CI 0.46 to 0.85), and routine occupations (OR = 0.62, 95% CI 0.39 to 0.98), and sixty-two-year-olds in lower supervisory and technical occupations (OR = 0.45, 95% CI 0.24 to 0.84) were less likely to report an increase in alcohol use compared to those in higher managerial positions (Supporting information, Tables S6 and S7). Finally, among thirty-year-olds, non-White ethnicity was associated with a 29% (OR = 0.71, 95% CI 0.55 to 0.93) reduction in the odds of reporting increased drinking (Supporting information, Table S4).

**Figure 1.**
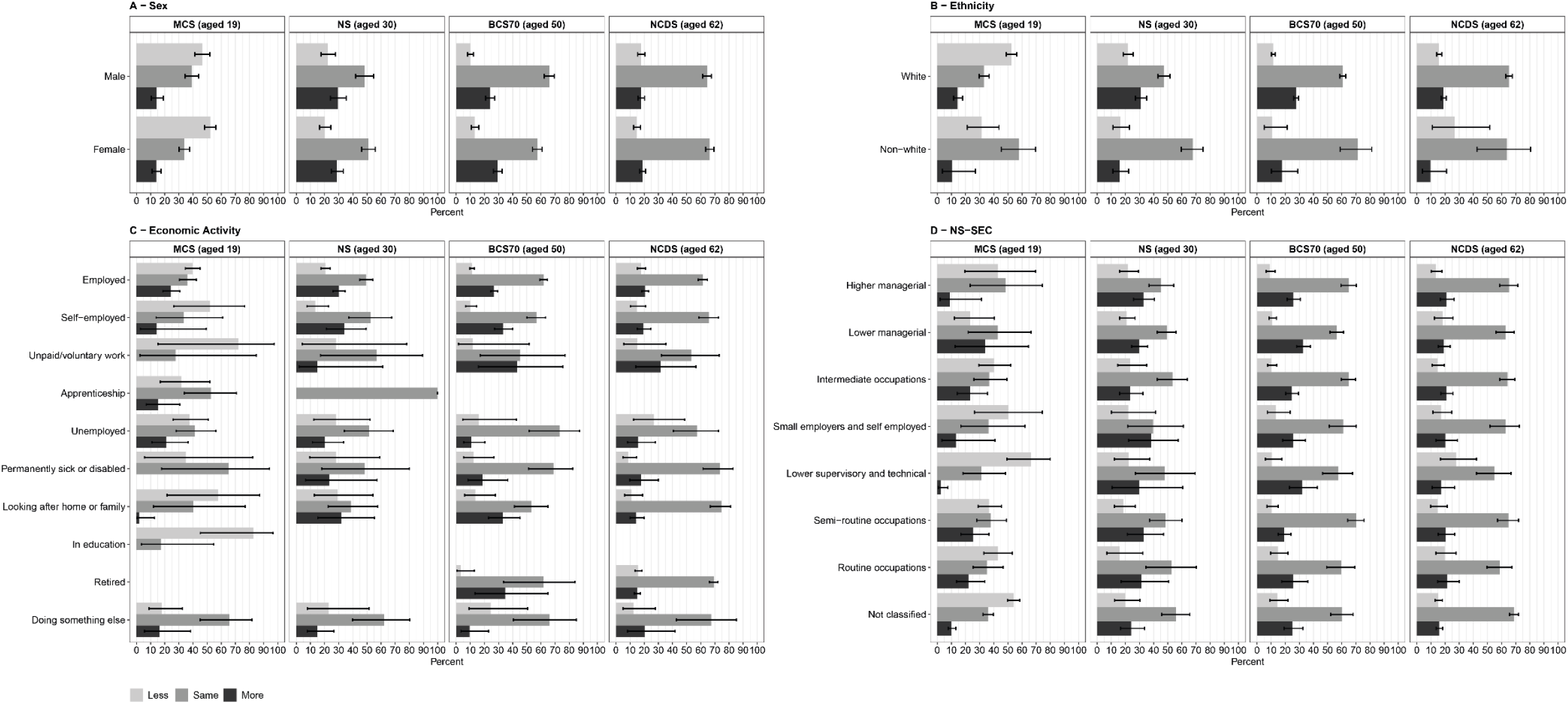
Change in alcohol use during the first wave (May 2020) of the COVID-19 pandemic in the UK, utilising data from four birth cohorts: The Millennium Cohort Study (*n* = 2,645), Next Steps (*n* = 1,907), the British Cohort Study (*n* = 4,223), and the National Child Development Study (*n* = *5*,178) by sex (panel A), ethnicity (panel B), economic activity during the pandemic (panel C), and National Statistics Socio-economic Class (panel D). Point estimates represent weighted percentages, error bars represent 95% confidence intervals.

### Risk of alcohol-related harm due to hazardous drinking during first lockdown

Most participants fell into the low-risk category regardless of age or sub-group membership since the start of the lockdown (Table 1, Figure 2). Approximately one-fifth of nineteen-year-olds, one-third of thirty-year-olds, and two-fifths of both fifty-year-olds and sixty-two-year-olds were at an increased risk of alcohol-related harm or worse. Of these, approximately 60.50% (95% CI 48.73 to 71.17) of nineteen-year-olds, 59.93% (95% CI 52.51 to 66.92) of thirty-year-olds, 68.11% (95% CI 63.14 to 72.71) of fifty-year-olds, and 69.28% (95% CI 49.96, 73.29) of sixty-two-year-olds reported an increase in alcohol use since the start of the pandemic.

**Figure 2.**
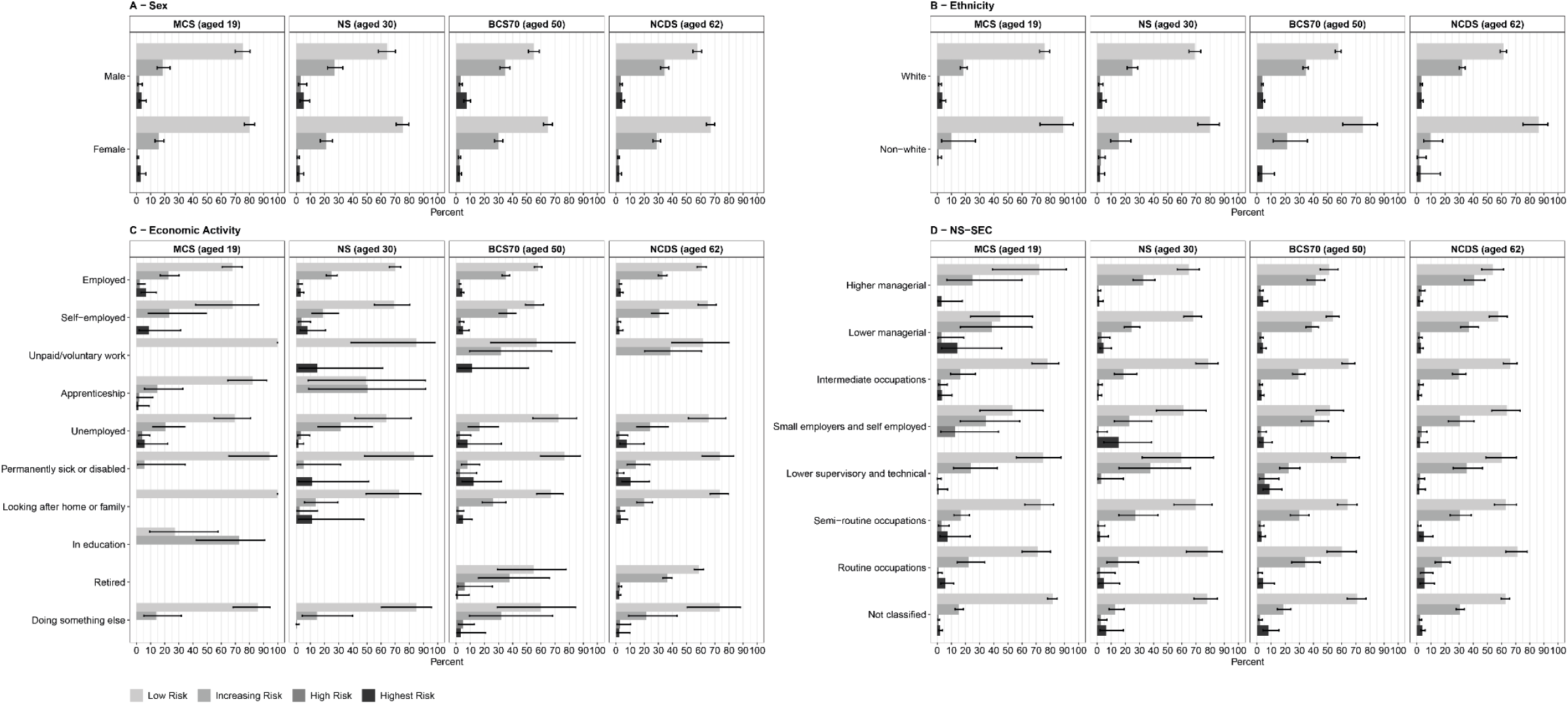
Risk of alcohol-related harm due to hazardous drinking during the first wave (May 2020) of the COVID-19 pandemic in the UK, utilising data from four birth cohorts: The Millennium Cohort Study (*n* = 2,645), Next Steps (*n* = 1,907), the British Cohort Study (*n* = 4,223), and the National Child Development Study (*n* = *5*,178) by sex (panel A), ethnicity (panel B), economic activity during the pandemic (panel C), and National Statistics Socio-economic Class (panel D). Point estimates represent weighted percentages, error bars represent 95% confidence intervals.

Figure 2 shows risk of alcohol-related harm due to hazardous drinking by sub-group. Among nineteen-year-olds, being employed or in education was associated with an increase in the odds of being more at risk of alcohol-related harm (Supporting information, Table S8). For thirty, fifty, and sixty-two-year-olds (Supporting information, Tables S9 – S11), being female and non-White ethnicity was associated with decreased odds of alcohol-related harm due to hazardous drinking. Finally, some effects were cohort specific. Being a permanently sick or disabled fifty-year-old was associated with a 76% (OR = 0.24, 95% CI 0.10 to 0.58) decrease in the odds of alcohol related harm compared to those who were employed (Supporting information, Table S10). Similarly, sixty-two-year-olds who worked in routine occupations (OR = 0.56, 95% CI 0.33 to 0.96) were less likely to drink hazardously (Supporting information, Table S11).

### Change in stress

Across all cohorts, most participants reported experiencing the same amount or less stress since the start of the pandemic (Table 1). Approximately two-fifths of nineteen-year-olds, half of thirty-year-olds, two-fifths of fifty-year-olds, and one-third of sixty-two-year-olds reported feeling more stressed. Of those, females were disproportionately affected (Figure 3). More specifically, among nineteen-year-olds, being female was associated with 1.54 (95% CI 1.08 to 2.20) times the odds of reporting an increase in stress (Table S12). For thirty-year-olds, being female was associated with 1.93 (95% CI 1.39 to 2.70) times the odds of reporting an increase in stress (Supporting information, Table S13). For fifty-year-olds, being female was associated with 1.62 (95% CI 1.37 to 1.92) times the odds of reporting an increase in stress (Supporting information, Table S14). For sixty-two-year-olds, being female was associated with 2.03 (95% CI 1.66 to 2.48) times the odds of reporting an increase in stress (Supporting information, Table S15). Additionally, for nineteen-year-olds being either self-employed (OR = 5.53, 95% CI 1.56 to 19.57) or unemployed (OR = 1.75, 95% CI 1.08 to 2.83) was associated with an increase in the odds of reporting an increase in stress (Supporting information, Table S12). Similarly, for thirty-year-olds, being unemployed (OR = 2.14, 95% CI 1.15 to 3.98) was also associated with an increase in the odds of reporting an increase in stress (Supporting information, Table S13).

**Figure 3.**
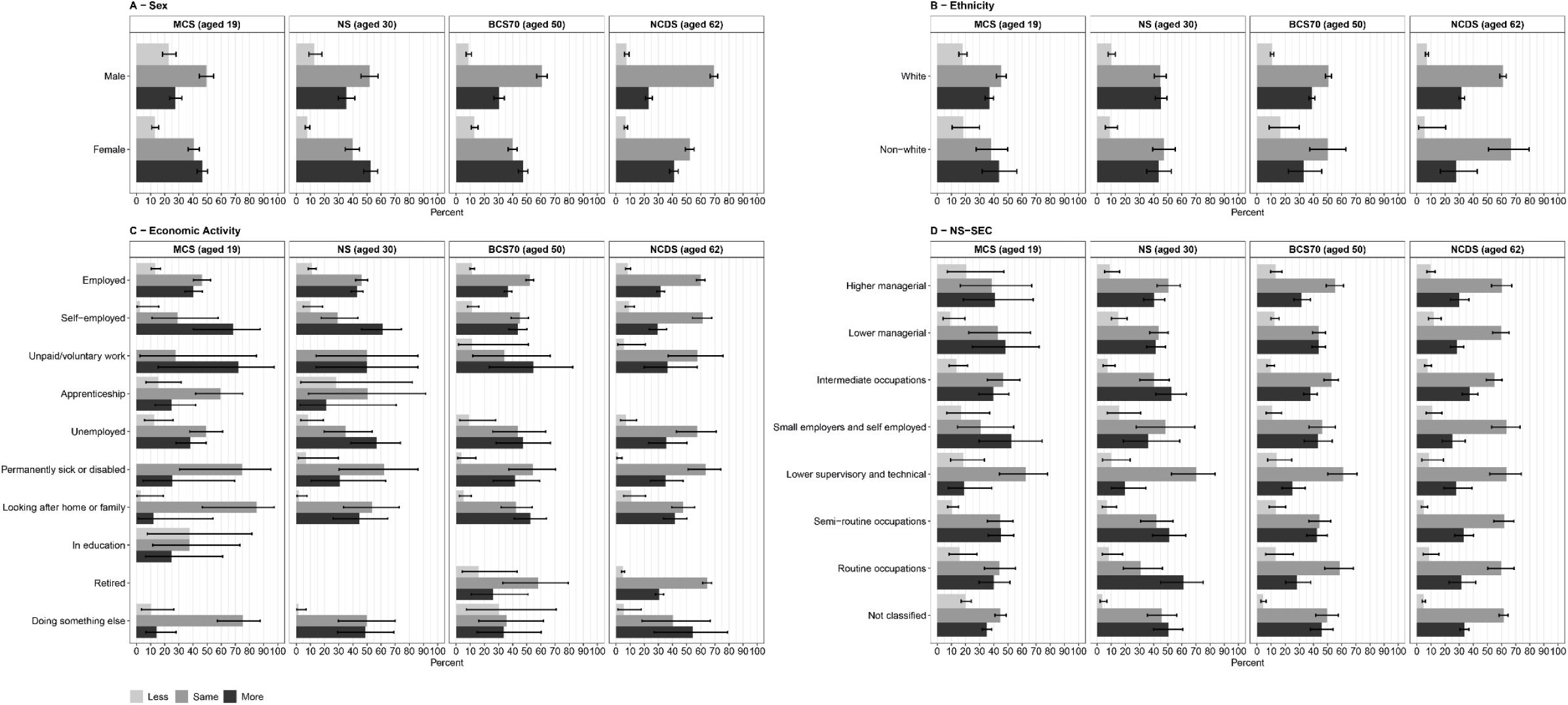
Change in perceived stress during the first wave (May 2020) of the COVID-19 pandemic in the UK, utilising data from four birth cohorts: The Millennium Cohort Study (*n* = 2,645), Next Steps (*n* = 1,907), the British Cohort Study (*n* = 4,223), and the National Child Development Study (*n* = *5*,178) by sex (panel A), ethnicity (panel B), economic activity during the pandemic (panel C), and National Statistics Socio-economic Class (panel D). Point estimates represent weighted percentages, error bars represent 95% confidence intervals.

### Associations between stress, inhibitory control, and drinking behaviour

#### Stress

After adjusting for potential confounders, thirty-year-olds who reported feeling more stressed since the start of lockdown were at 3.77 (95% CI 1.15 to 12.28) times greater odds of being at increasing, high, or highest (versus low) risk of alcohol-related harm, compared to those that reported feeling no change in stress (Table 2). There was no evidence to suggest that this effect was present in other cohorts.

**Table 2.**
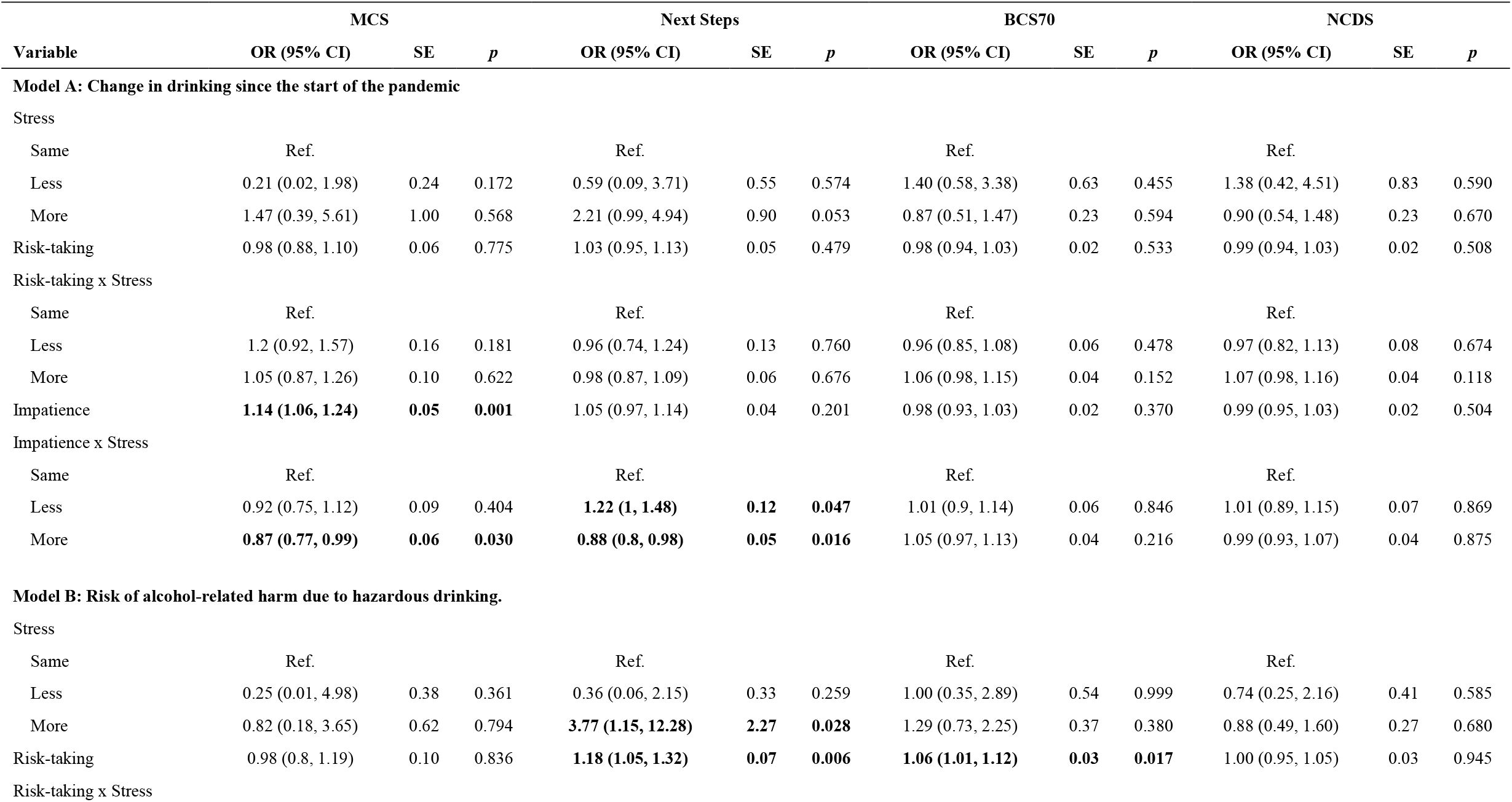
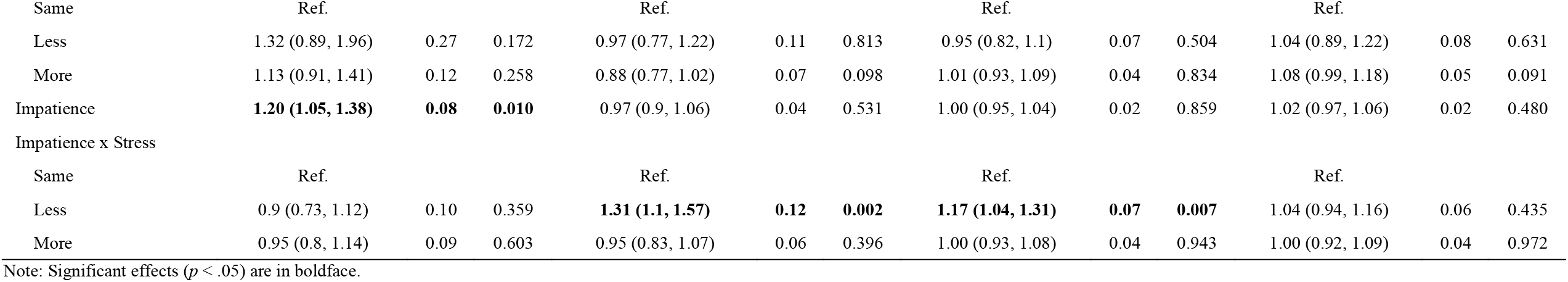
Summary of the final ordinal regression models predicting change in drinking since the start of the pandemic (model A) and risk of alcohol–related harm due to hazardous drinking during the pandemic (model B), adjusting for sex, ethnicity, economic activity during the pandemic, and social class prior to the pandemic.

|  | MCS |  |  | Next Steps |  |  | BCS70 |  |  | NCDS |  |  |
| --- | --- | --- | --- | --- | --- | --- | --- | --- | --- | --- | --- | --- |
| Variable | OR (95% CI) | SE | p | OR (95% CI) | SE | p | OR (95% CI) | SE | p | OR (95% CI) | SE | p |
| Model A: Change in drinking since the start of the pandemic |  |  |  |  |  |  |  |  |  |  |  |  |
| Stress |  |  |  |  |  |  |  |  |  |  |  |  |
| Same | Ref. |  |  | Ref. |  |  | Ref. |  |  | Ref. |  |  |
| Less | 0.21 (0.02, 1.98) | 0.24 | 0.172 | 0.59 (0.09, 3.71) | 0.55 | 0.574 | 1.40 (0.58, 3.38) | 0.63 | 0.455 | 1.38 (0.42, 4.51) | 0.83 | 0.590 |
| More | 1.47 (0.39, 5.61) | 1.00 | 0.568 | 2.21 (0.99, 4.94) | 0.90 | 0.053 | 0.87 (0.51, 1.47) | 0.23 | 0.594 | 0.90 (0.54, 1.48) | 0.23 | 0.670 |
| Risk-taking | 0.98 (0.88, 1.10) | 0.06 | 0.775 | 1.03 (0.95, 1.13) | 0.05 | 0.479 | 0.98 (0.94, 1.03) | 0.02 | 0.533 | 0.99 (0.94, 1.03) | 0.02 | 0.508 |
| Risk-taking x Stress |  |  |  |  |  |  |  |  |  |  |  |  |
| Same | Ref. |  |  | Ref. |  |  | Ref. |  |  | Ref. |  |  |
| Less | 1.2 (0.92, 1.57) | 0.16 | 0.181 | 0.96 (0.74, 1.24) | 0.13 | 0.760 | 0.96 (0.85, 1.08) | 0.06 | 0.478 | 0.97 (0.82, 1.13) | 0.08 | 0.674 |
| More | 1.05 (0.87, 1.26) | 0.10 | 0.622 | 0.98 (0.87, 1.09) | 0.06 | 0.676 | 1.06 (0.98, 1.15) | 0.04 | 0.152 | 1.07 (0.98, 1.16) | 0.04 | 0.118 |
| Impatience | **1.14 (1.06, 1.24)** | **0.05** | **0.001** | 1.05 (0.97, 1.14) | 0.04 | 0.201 | 0.98 (0.93, 1.03) | 0.02 | 0.370 | 0.99 (0.95, 1.03) | 0.02 | 0.504 |
| Impatience x Stress |  |  |  |  |  |  |  |  |  |  |  |  |
| Same | Ref. |  |  | Ref. |  |  | Ref. |  |  | Ref. |  |  |
| Less | 0.92 (0.75, 1.12) | 0.09 | 0.404 | **1.22 (1, 1.48)** | **0.12** | **0.047** | 1.01 (0.9, 1.14) | 0.06 | 0.846 | 1.01 (0.89, 1.15) | 0.07 | 0.869 |
| More | **0.87 (0.77, 0.99)** | **0.06** | **0.030** | **0.88 (0.8, 0.98)** | **0.05** | **0.016** | 1.05 (0.97, 1.13) | 0.04 | 0.216 | 0.99 (0.93, 1.07) | 0.04 | 0.875 |
| Model B: Risk of alcohol-related harm due to hazardous drinking. |  |  |  |  |  |  |  |  |  |  |  |  |
| Stress |  |  |  |  |  |  |  |  |  |  |  |  |
| Same | Ref. |  |  | Ref. |  |  | Ref. |  |  | Ref. |  |  |
| Less | 0.25 (0.01, 4.98) | 0.38 | 0.361 | 0.36 (0.06, 2.15) | 0.33 | 0.259 | 1.00 (0.35, 2.89) | 0.54 | 0.999 | 0.74 (0.25, 2.16) | 0.41 | 0.585 |
| More | 0.82 (0.18, 3.65) | 0.62 | 0.794 | **3.77 (1.15, 12.28)** | **2.27** | **0.028** | 1.29 (0.73, 2.25) | 0.37 | 0.380 | 0.88 (0.49, 1.60) | 0.27 | 0.680 |
| Risk-taking | 0.98 (0.8, 1.19) | 0.10 | 0.836 | **1.18 (1.05, 1.32)** | **0.07** | **0.006** | **1.06 (1.01, 1.12)** | **0.03** | **0.017** | 1.00 (0.95, 1.05) | 0.03 | 0.945 |
| Risk-taking x Stress |  |  |  |  |  |  |  |  |  |  |  |  |

| Same | Ref. |  |  | Ref. |  |  | Ref. |  |  | Ref. |  |  |
| --- | --- | --- | --- | --- | --- | --- | --- | --- | --- | --- | --- | --- |
| Less | 1.32 (0.89, 1.96) | 0.27 | 0.172 | 0.97 (0.77, 1.22) | 0.11 | 0.813 | 0.95 (0.82, 1.1) | 0.07 | 0.504 | 1.04 (0.89, 1.22) | 0.08 | 0.631 |
| More | 1.13 (0.91, 1.41) | 0.12 | 0.258 | 0.88 (0.77, 1.02) | 0.07 | 0.098 | 1.01 (0.93, 1.09) | 0.04 | 0.834 | 1.08 (0.99, 1.18) | 0.05 | 0.091 |
| Impatience | **1.20 (1.05, 1.38)** | **0.08** | **0.010** | 0.97 (0.9, 1.06) | 0.04 | 0.531 | 1.00 (0.95, 1.04) | 0.02 | 0.859 | 1.02 (0.97, 1.06) | 0.02 | 0.480 |
| Impatience x Stress |  |  |  |  |  |  |  |  |  |  |  |  |
| Same | Ref. |  |  | Ref. |  |  | Ref. |  |  | Ref. |  |  |
| Less | 0.9 (0.73, 1.12) | 0.10 | 0.359 | **1.31 (1.1, 1.57)** | **0.12** | **0.002** | **1.17 (1.04, 1.31)** | **0.07** | **0.007** | 1.04 (0.94, 1.16) | 0.06 | 0.435 |
| More | 0.95 (0.8, 1.14) | 0.09 | 0.603 | 0.95 (0.83, 1.07) | 0.06 | 0.396 | 1.00 (0.93, 1.08) | 0.04 | 0.943 | 1.00 (0.92, 1.09) | 0.04 | 0.972 |

Note: Significant effects ( $p < .05$ ) are in boldface.

#### Impatience

Among nineteen-year-olds, a one unit increase in impatience was associated with 1.14 (95% CI 1.06 to 1.24) times the odds of reporting an increase in alcohol use, and 1.20 (OR = 1.20, 95% CI 1.05 to 1.38) times the odds of alcohol-related harm due to hazardous drinking after controlling for potential confounders (Table 2). There was no evidence to suggest that this effect was present in other cohorts.

#### Risk–taking

After controlling for potential confounders, a one unit increase in risk-taking was associated with 1.18 (95% CI 1.05 to 1.32) times the odds of alcohol-related harm among thirty-year-olds (Table 2). Similarly, for fifty-year-olds, a one unit increase in risk-taking was associated with 1.06 (95% CI 1.01 to 1.12) times the odds of alcohol-related harm. This effect was not observed in other cohorts (Table 2).

#### Stress x personality interactions

There was evidence to suggest that, after controlling for potential confounders, individuals who were more impatient and *<u>less stressed</u>* tended to drink more and be at a greater risk of alcohol-related harm (Table 2). Specifically, for thirty-year-olds, a one unit increase in impatience was associated with a 22% (OR = 1.22, 95% CI 1.00 to 1.48) increase in the odds of reporting an increase in alcohol use among those who reported feeling less stressed, and a 12% (OR = 0.88, 95% CI 0.80 to 0.98) decrease in the odds of reporting an increase in alcohol use among those who reported feeling more stressed. Similarly, among nineteen-year-olds that reported feeling more stressed, a one unit increase in impatience was associated with a 13% (OR = 0.87, 95% CI 0.77 to 0.99) decrease in the odds of reporting an increase in alcohol use. In terms of risk of alcohol-related harm, for both thirty-year-olds (OR = 1.31, 95% CI 1.10 to 1.57) and fifty-year-olds (OR = 1.17, 95% CI 1.04 to 1.31), reporting feeling less stressed was associated with an increase in the odds of being at an increased risk of alcohol-related harm or worse. No stress x personality interactions were observed in the sixty-two-year-old cohort. No stress x risk-taking interactions were observed in any group.

## Discussion

The present study utilised data from four nationally representative British birth cohorts to explore changes in alcohol use behaviour and stress since the start of the COVID-19 outbreak, during the first national lockdown in May 2020. Across all age-groups (cohorts), we found evidence to suggest that most respondents drank the same amount or less since the start of the pandemic. However, between approximately fourteen and thirty percent of respondents reported drinking more depending on age. Of these, thirty-year-olds and fifty-year-olds were most likely to report an increase in drinking. This supports recent emerging evidence which suggests that between one-fifth and one-third of individuals in the UK reported drinking more during the first wave of the pandemic [46–48]. Further, between twenty and forty percent of participants drank at levels of increasing risk of alcohol-related harm or worse, depending on age, with older participants displaying the greatest levels of risk due to alcohol misuse. Of these, approximately sixty percent of both nineteen-year-olds and thirty-year-olds, and seventy percent of both fifty-year-olds and sixty-two-year-olds reported drinking more since the start of the pandemic. Provisional data from the Office for National Statistics data suggests that alcohol-related deaths reached a 20-year high between quarter one (January to March) and quarter three (July to September) of 2020; with significant increases in mortality among those aged between thirty and forty-nine in quarter two and forty to sixty-nine in quarter three [49]. These data add concerning weight to our findings of the higher rates of harmful drinking in these age groups, supporting the public health concerns attributable to excess alcohol use in some at-risk individuals during lockdown [4–6]. The increase in alcohol-related deaths could be, at least partly, attributable to changes in mental health service provision during the pandemic and therefore increased psychological distress on top of that directly associated with stay-at-home orders [50,51].

Similar to changes in drinking behaviour, most participants reported experiencing the same amount or less stress since the start of the pandemic. Nevertheless, between approximately thirty and forty-five percent of respondents reported an increase in their stress level. Of these, thirty-year-olds seemed to be most affected as more respondents from this group reported increased stress compared to the other cohorts. This group also had the highest proportion of individuals that reported increased alcohol use and there was evidence of an association between stress and hazardous drinking here too. Analogous to this finding, previous research suggests that the Millennial generation struggle with stress management considerably more than previous generations [52]. Similarly, recent data from the UK Household Longitudinal Survey [53] suggests that young individuals have seen larger declines in well–being during the first lockdown. Surprisingly, despite the well-established link between substance use and stress [17,18], a main effect of stress was not observed in any other group. However, in all cohorts, being female was associated with an increased likelihood of reporting heightened stress; an effect which has consistently been reported elsewhere [48,53,54]. This may be due to (for example) an increased risk of psychiatric symptoms prior to, and after, suffering with COVID-19; an increased risk of domestic violence; and a disproportionate responsibility for domestic tasks including caring for family members [55]. In terms of drinking, our results suggest that for the fifty- and sixty-two-year-olds cohorts, being female was associated with an approximate twenty-five percent increase in the odds of reporting an increased alcohol use. Interestingly, however, across all cohorts, except the nineteen-year-olds, being female was associated with around a forty percent reduction in the odds of alcohol-related harm due to hazardous drinking.

Several sociodemographic characteristics were related to change in both stress and alcohol use behaviour. For instance, in all but the oldest cohorts, employment was related to reporting increased alcohol use; and in the youngest cohort, both being employed or in-education was associated with an increased likelihood of hazardous drinking and subsequent alcohol-related harm. Similarly, among fifty-year-olds those in higher managerial positions were more likely to report increased alcohol use. Meanwhile, for those aged sixty-two, higher managerial positions were associated with an increased risk of alcohol related harm due to hazardous drinking. As off-premises alcohol consumption has been classified as ‘essential’ by the UK government [56], this association is likely related to the physical and financial availability of alcohol [5,57]. In other words, those that are employed and/or high earners will generally be able to (financially) afford to drink more. Regarding changes in stress, unemployment was related to an increased likelihood of reporting heightened stress among both nineteen- and thirty-year-olds. Also, self-employed nineteen-year-olds were more likely to report increased stress. Again, this was most likely associated with financial stability. For instance, many people who rely on state welfare have been receiving Universal Credit which has been shown to be associated with psychological distress [58], and recent research has shown that self–employed people have suffered a large and disproportionate reduction in income during the pandemic [59]. Finally, in all but the youngest cohorts, there was evidence to suggest that non-White ethnicity was associated with a decreased likelihood of alcohol-related harm due to hazardous drinking; and among thirty-year-olds non-White ethnicity was associated with a decreased likelihood of reporting and increase in alcohol use. This was unsurprising considering that results from several papers suggest that being White is a risk-factor for alcohol use and misuse [60–62].

Self-reported inhibitory control, and in some cases, a complex interaction between stress and personality were related to alcohol use and hazardous drinking during the lockdown. For example, in thirty– and fifty-year–olds, risk-taking personality was associated with an increased propensity to consume more alcohol and to have higher hazardous drinking scores. This corresponds to a large volume of literature which associates poor inhibitory control with substance misuse [7,8,11–16].

Moreover, the majority of nineteen–year–olds reported drinking less since the start of the pandemic. This was unsurprising considering the recent evidence of the ‘devaluation of alcohol’ among Generation Z [63]. This finding may also have been driven by the closure of on–trade drinking locations since drinking at venues such as pubs and bars is more common among young people [64], and reduced exposure to environments related with alcohol consumption has been associated with a reduction in drinking among young individuals during the pandemic [65]. However, critically, for nineteen-year-olds, impatience was related to increased alcohol use and risk of alcohol-related harm due to hazardous drinking during the pandemic. This group also had the highest levels of impatience across all cohorts. Taken together, these findings raise a concern about the potential for adults who have poor inhibitory control to be at particular risk of an escalation of alcohol misuse following the pandemic situation.

It is clear from previous research that there is an interaction between stress and personality factors that influences drinking behaviour. For example, people who experience acute stress show increases in craving for, and consumption of, alcohol [24,25]. Here, counterintuitively, we found that greater impatience and decreased stress was associated with increased alcohol use among thirty-year-olds and an increased hazardous drinking among both thirty-year-olds and fifty-year-olds. Similarly, among nineteen-and thirty-year-olds, those that rated themselves as more impatient and experienced increased stress were less likely to report increased alcohol consumption. As ‘drinking to cope’ was a prominent feature related to alcohol use during lockdown in the USA [66], it may also be the case here. For instance, individuals with poor inhibitory control tend to use alcohol as a method of dealing with stress [67,68]. Therefore, these individuals may have reduced stress levels due to their reported increased alcohol use. Alternatively, as the physiological response to long–term (chronic) and short– term (acute) stress differs [69], it may be that the interaction between inhibitory control and chronic stress also differs. Therefore, future research should endeavour to investigate the impact of the interaction between different types of stress and inhibitory control in the context of alcohol use.

### Limitations

We acknowledge several limitations in our study. First, the survey was designed to capture information across several domains other than those relevant here. Therefore, to mitigate known issues related to respondent burden (e.g., satisficing), brevity was prioritised, which inevitably resulted in less detail than may be ideal in some of the measures used. For instance, single-item measures were used to assess risk-taking, impatience, and stress which may fail adequately to capture the full scope of these constructs (i.e., these measures may suffer from reduced content validity). This increases the uncertainty surrounding estimates calculated using these measures. Therefore, the use of single–item measures may also inflate standard errors and risk for type II error. Some of this potential error is offset by our large sample size; however, we found some effects that were not statistically significant despite relatively large effect sizes (e.g., among thirty-year-olds that reported increased stress, OR = 2.21, 95% CI 0.99 to 4.94). Second, there may be individual differences in the way each question was interpreted. For instance, feelings of stress are subjective and vary between–individuals [70]. Therefore, while some may find the pandemic and related period of social isolation as extremely stressful, others will find lockdown less stressful than pre–pandemic life. This may offer another explanation for why some that reported poor inhibitory control and lower levels of stress also reported increased alcohol use. Third, there is no way to independently verify self-report drinking; it is well-known that people under-estimate their alcohol consumption when asked on questionnaires due to social desirability bias, and often a lack of detailed memory of drinking episodes [71]. It may, therefore, be that our data under-represent the true extent of drinking during the pandemic. Finally, the longitudinal nature of birth cohort data allows for attrition-related bias to be minimised using sample weights calculated by the CLS team [27]. However, there is a possibility that unobserved predictors of missing data may still influence results.

## Conclusions

In conclusion, we aimed to explore factors that influenced changes in alcohol use behaviour during the first COVID-19 lockdown in the UK, particularly concentrating on self-report stress and personality characteristics (risk-taking and impatience). We found that although most respondents drank either the same amount or less than prior to the pandemic, a significant minority, particularly of thirty- and fifty-year olds, drank more; often in amounts which could be classified hazardous, thus increasing their risk of potential alcohol-related harm. We also found that increases in drinking hazardously were predicted by personality (risk-taking, impatience) and environment (stress), although this was age specific. When considered in combination with recent data on alcohol-related deaths in the UK during the first three quarters of 2020, our findings suggest that hazardous drinking in a minority was strongly influenced by the pandemic and propose that this may be influenced by a combination of stress and personality factors, but also likely due to the availability of alcohol and inaccessible mental health services. We suggest that in future lockdowns, the government and public health officials pay particular attention to at-risk individuals, in terms of service provision, and consider critically the ‘essential’ nature of off-premises alcohol sales.

## Data Availability

All data used in this study are available from the UK Data Service Website (https://ukdataservice.ac.uk/) under the "Safeguarded" data access policy. The primary dataset used is referenced under study number 8658. Ethnicity data associated with the MCS, Next Steps, BCS70, and NCDS cohorts are referenced under study numbers 6073, 5545, 5558, and 5565 respectively. All analysis code is available on the Open Science Framework website (https://osf.io/wf2rj/).

## Acknowledgements

We would like to thank Centre for Longitudinal Studies team at UCL for collecting the data analysed here and for making the data available through the UK Data Service. The Centre for Longitudinal Studies receive funding from the ESRC, the MRC, the NIH, Welcome, the British Heart Foundation, the Academy of Medical Science, the Health Foundation, and the UCL Institute of Education (https://cls.ucl.ac.uk/about-2/our-funders/). JMC is funded by an ESRC Doctoral Training Partnership grant (ES/P000673/1). MOP receives funding from the Foundation for Liver Research. The funders had no role in study design, data analysis, data interpretation, writing of the report, or the decision to submit for publication. We would also like to thank all of the participants that completed the survey for their cooperation.

## Author Contributions

Contributions are indicated using the CRediT (Contribution Roles Taxonomy).

**James Clay**: Conceptualisation; methodology; validation; formal analysis; investigation; data curation; writing – original draft; visualisation; project administration; funding acquisition.

**Lorenzo Stafford**: Conceptualisation; methodology; validation; writing – review and editing; supervision; funding acquisition.

**Matthew Parker**: Conceptualisation; methodology; validation; formal analysis; investigation; writing – review and editing; supervision; project administration; funding acquisition.

## Notes

### Competing Interest Statement

The authors have declared no competing interest.

### Funding Statement

JMC is funded by an ESRC Doctoral Training Partnership grant. MOP receives funding from the Foundation for Liver Research. The funders had no role in study design, data analysis, data interpretation, writing of the report, or the decision to submit for publication. The UCL Centre for Longitudinal Studies, who administered the survey, played no role in the analysis of the data, preparation of the manuscript, or decision to submit the manuscript for publication. All authors had full access to all data in the study. All authors were responsible for the decision to submit for publication.

### Author Declarations

The data used in this work were from the UK Data Service. The registration for the access to the data from UK Data Service was done via University of Portsmouth. Access to the data was given by UK Data Service with the Project Number: 197137. As a secondary analysis was carried out, full ethical review was not required. Thus, the University of Portsmouth Ethics Screening Tool was used for assessment (reference number: ETHICS-10155).

### Summary of Updates

Updated manuscript and supplementary materials.

