## Supporting Information for "Associations between inhibitory control, stress, and alcohol (mis)use during the first wave of the COVID-19 pandemic in the UK: a national cross-sectional study utilising data from four birth cohorts"

#### Supplementary Methods

**Figure S1.** Study sample overview. MCS = Millennium Cohort Study (born 2000 – 02); NSHD = MRC National Survey of Health and Development (born 1946); NCDS = 1958 National Child Development Study; BCS70 = 1970 British Cohort Study; the Next Steps cohort were born 1989 – 90. Opted out = Those who requested not to be contacted further via phone, email, or by clicking the "opt-out" button, which was included in the invitation email. Responded = Anyone who completed the first block of the questionnaire. Data from Brown et al., 2020 [1].

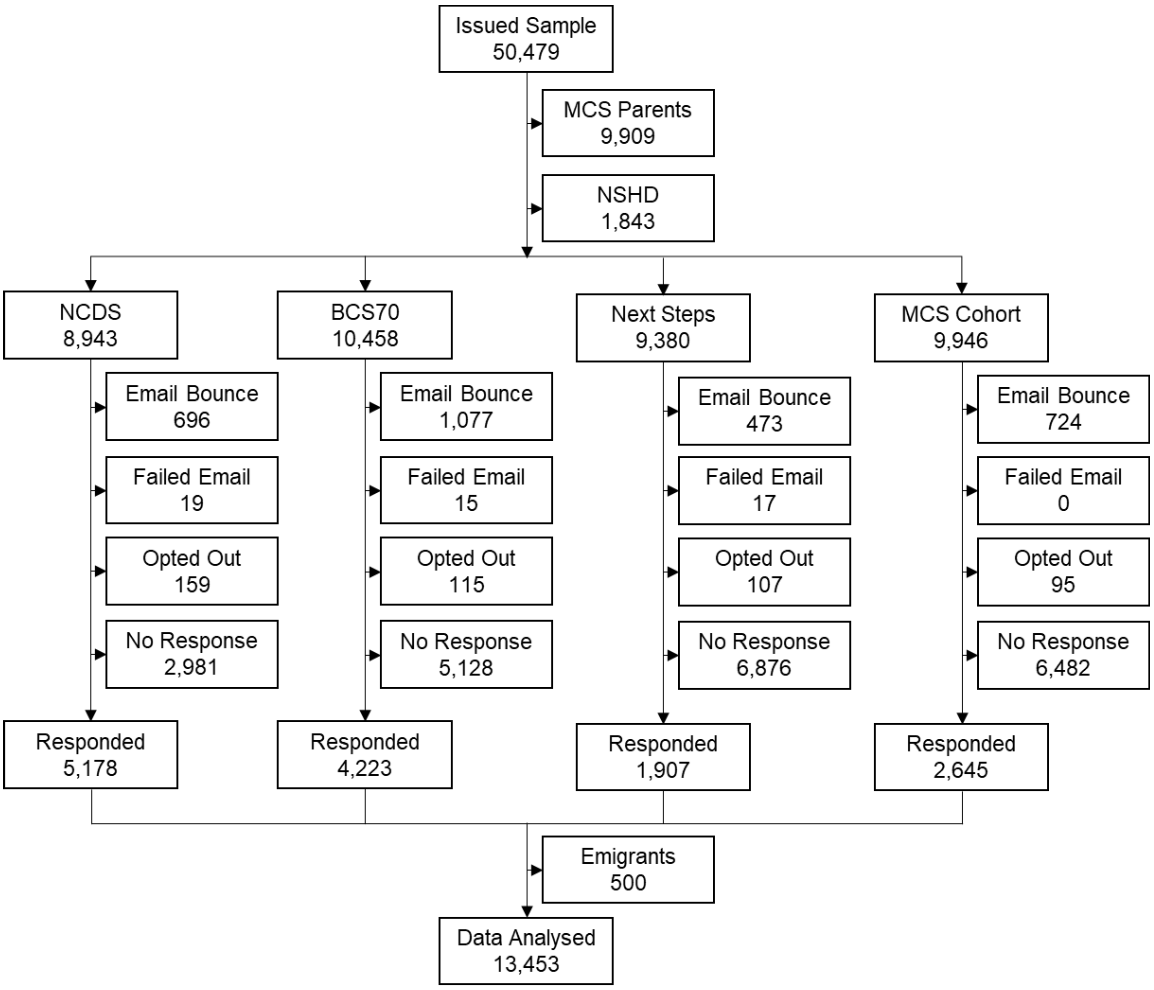

**Table S1.** Linear regression models used to assess the association between stress and PHQ-4 score.

| | MCS | | | | | | Next Steps | | | | | | BCS70 | | | | | | NCDS | | | | | |
|---|---|---|---|---|---|---|---|---|---|---|---|---|---|---|---|---|---|---|---|---|---|---|---|---|
| **Variable** | **Coef.** | **LL** | **UL** | **SE** | ***t*** | ***p*** | **Coef.** | **LL** | **UL** | **SE** | ***t*** | ***p*** | **Coef.** | **LL** | **UL** | **SE** | ***t*** | ***p*** | **Coef.** | **LL** | **UL** | **SE** | ***t*** | ***p*** |
| **Unadjusted model** | | | | | | | | | | | | | | | | | | | | | | | | |
| Stress | | | | | | | | | | | | | | | | | | | | | | | | |
| Same - no change | Ref | | | | | | Ref | | | | | | Ref | | | | | | Ref | | | | | |
| Less than before | -0.95 | -1.40 | -0.51 | 0.23 | -4.18 | **< .001** | -0.40 | -0.80 | -0.01 | 0.20 | -2.00 | **0.046** | 0.20 | -0.16 | 0.57 | 0.19 | 1.10 | 0.272 | 0.00 | -0.28 | 0.29 | 0.15 | 0.02 | 0.983 |
| More than before | 2.45 | 2.02 | 2.88 | 0.22 | 11.21 | **< .001** | 2.60 | 2.17 | 3.04 | 0.22 | 11.72 | **< .001** | 2.16 | 1.78 | 2.54 | 0.19 | 11.09 | **< .001** | 2.16 | 1.89 | 2.44 | 0.14 | 15.48 | **< .001** |
| **Adjusted model** | | | | | | | | | | | | | | | | | | | | | | | | |
| Stress | | | | | | | | | | | | | | | | | | | | | | | | |
| Same - no change | Ref | | | | | | Ref | | | | | | Ref | | | | | | Ref | | | | | |
| Less than before | -0.41 | -0.94 | 0.12 | 0.27 | -1.52 | 0.13 | -0.21 | -0.62 | 0.19 | 0.20 | -1.04 | 0.297 | 0.28 | -0.03 | 0.59 | 0.16 | 1.76 | 0.078 | 0.06 | -0.20 | 0.32 | 0.13 | 0.44 | 0.657 |
| More than before | 2.68 | 1.97 | 3.39 | 0.36 | 7.41 | **< .001** | 2.49 | 2.07 | 2.91 | 0.21 | 11.65 | **< .001** | 1.96 | 1.72 | 2.19 | 0.12 | 16.47 | **< .001** | 1.95 | 1.68 | 2.23 | 0.14 | 13.87 | **< .001** |
| Sex | | | | | | | | | | | | | | | | | | | | | | | | |
| Male | Ref | | | | | | Ref | | | | | | Ref | | | | | | Ref | | | | | |
| Female | 0.68 | 0.05 | 1.32 | 0.32 | 2.13 | **0.03** | 0.50 | 0.09 | 0.91 | 0.21 | 2.42 | **0.016** | 0.01 | -0.22 | 0.24 | 0.12 | 0.07 | 0.941 | 0.27 | 0.02 | 0.52 | 0.13 | 2.12 | **0.034** |
| Ethnicity | | | | | | | | | | | | | | | | | | | | | | | | |
| White | Ref | | | | | | Ref | | | | | | Ref | | | | | | Ref | | | | | |
| Non-white | 0.04 | -0.85 | 0.94 | 0.45 | 0.09 | 0.93 | 0.05 | -0.37 | 0.47 | 0.21 | 0.24 | 0.809 | -0.22 | -0.58 | 0.13 | 0.18 | -1.24 | 0.213 | 0.25 | -0.31 | 0.81 | 0.29 | 0.87 | 0.385 |
| NS-SEC 2010 analytical classes | | | | | | | | | | | | | | | | | | | | | | | | |
| Higher managerial | Ref | | | | | | Ref | | | | | | Ref | | | | | | Ref | | | | | |
| Lower managerial | 0.47 | -1.39 | 2.33 | 0.94 | 0.50 | 0.62 | -0.18 | -0.73 | 0.37 | 0.28 | -0.64 | 0.525 | 0.00 | -0.25 | 0.25 | 0.13 | 0.03 | 0.976 | 0.08 | -0.18 | 0.34 | 0.13 | 0.61 | 0.544 |
| Intermediate occupations | 0.28 | -1.22 | 1.78 | 0.76 | 0.37 | 0.71 | 0.53 | -0.23 | 1.30 | 0.39 | 1.38 | 0.169 | 0.31 | 0.02 | 0.60 | 0.15 | 2.08 | **0.038** | 0.19 | -0.09 | 0.47 | 0.14 | 1.33 | 0.183 |
| Small employer and self-employed | 0.25 | -1.98 | 2.48 | 1.13 | 0.22 | 0.83 | 0.43 | -0.56 | 1.43 | 0.51 | 0.85 | 0.394 | -0.17 | -0.52 | 0.18 | 0.18 | -0.95 | 0.343 | 0.18 | -0.19 | 0.54 | 0.19 | 0.95 | 0.344 |
| Lower supervisory and technical | 0.04 | -1.71 | 1.80 | 0.89 | 0.05 | 0.96 | -0.37 | -0.95 | 0.21 | 0.29 | -1.25 | 0.212 | 0.57 | -0.02 | 1.17 | 0.30 | 1.89 | 0.059 | -0.11 | -0.59 | 0.37 | 0.25 | -0.45 | 0.655 |
| Semi-routine occupations | 0.38 | -1.22 | 1.98 | 0.81 | 0.47 | 0.64 | 0.57 | -0.24 | 1.39 | 0.41 | 1.38 | 0.169 | 0.53 | 0.10 | 0.95 | 0.22 | 2.42 | 0.016 | 0.49 | 0.04 | 0.94 | 0.23 | 2.15 | **0.032** |
| Routine occupations | 1.02 | -0.67 | 2.71 | 0.86 | 1.19 | 0.24 | -0.02 | -1.13 | 1.09 | 0.56 | -0.04 | 0.970 | 0.40 | -0.07 | 0.87 | 0.24 | 1.67 | 0.095 | 0.41 | -0.10 | 0.91 | 0.26 | 1.57 | 0.117 |
| Uncategorised | 1.30 | -0.35 | 2.96 | 0.84 | 1.55 | 0.12 | 0.53 | -0.59 | 1.65 | 0.57 | 0.93 | 0.352 | 0.46 | 0.03 | 0.88 | 0.22 | 2.11 | **0.035** | 0.38 | -0.11 | 0.86 | 0.25 | 1.51 | 0.131 |
| Economic activity | | | | | | | | | | | | | | | | | | | | | | | | |
| Employed | Ref | | | | | | Ref | | | | | | Ref | | | | | | Ref | | | | | |
| Self-employed | -1.00 | -3.04 | 1.04 | 1.04 | -0.96 | 0.34 | -0.37 | -1.14 | 0.41 | 0.40 | -0.92 | 0.356 | -0.22 | -0.47 | 0.03 | 0.13 | -1.73 | 0.084 | 0.05 | -0.31 | 0.41 | 0.18 | 0.26 | 0.795 |
| In unpaid/voluntary work | 2.02 | -0.76 | 4.80 | 1.41 | 1.43 | 0.15 | -0.41 | -2.82 | 2.00 | 1.23 | -0.33 | 0.738 | -1.39 | -1.95 | -0.84 | 0.28 | -4.90 | **< .001** | -0.07 | -1.21 | 1.07 | 0.58 | -0.12 | 0.908 |
| Apprenticeship | -0.39 | -1.56 | 0.79 | 0.60 | -0.65 | 0.52 | 0.44 | -1.05 | 1.94 | 0.76 | 0.58 | 0.559 | - | - | - | - | - | - | - | - | - | - | - | - |
| Unemployed | 0.16 | -0.73 | 1.04 | 0.45 | 0.35 | 0.73 | 1.39 | -0.01 | 2.78 | 0.71 | 1.95 | 0.051 | 0.67 | -0.22 | 1.56 | 0.45 | 1.47 | 0.141 | 0.26 | -0.34 | 0.87 | 0.31 | 0.86 | 0.392 |
| Permanently sick or disabled | 2.86 | -2.31 | 8.02 | 2.62 | 1.09 | 0.28 | 4.28 | 1.63 | 6.93 | 1.35 | 3.17 | **0.002** | 3.65 | 2.31 | 5.00 | 0.69 | 5.32 | **< .001** | 3.32 | 1.87 | 4.77 | 0.74 | 4.50 | **< .001** |
| Looking after home or family | -1.30 | -2.68 | 0.08 | 0.70 | -1.86 | 0.07 | 0.90 | -0.58 | 2.39 | 0.75 | 1.20 | 0.231 | -0.03 | -0.53 | 0.46 | 0.25 | -0.13 | 0.894 | -0.26 | -0.86 | 0.35 | 0.31 | -0.84 | 0.401 |
| In education | 0.34 | -1.27 | 1.96 | 0.82 | 0.42 | 0.68 | - | - | - | - | - | - | - | - | - | - | - | - | - | - | - | - | - | - |
| Retired | - | - | - | - | - | - | - | - | - | - | - | - | -0.61 | -1.16 | -0.06 | 0.28 | -2.17 | **0.030** | -0.25 | -0.75 | 0.25 | 0.25 | -0.98 | 0.326 |
| Uncategorised | -1.12 | -2.39 | 0.15 | 0.64 | -1.73 | 0.08 | -0.01 | -1.26 | 1.25 | 0.64 | -0.01 | 0.992 | 0.07 | -1.36 | 1.49 | 0.73 | 0.09 | 0.925 | 0.46 | -0.67 | 1.59 | 0.58 | 0.80 | 0.422 |

Note: NS-SEC = National Statistics Socio-economic class prior to the outbreak. Economic activity reflects activity during the pandemic.

**Table S2.** Associations between single-item assessments, and behavioural assessments of inhibitory control.

| | Spearman's Correlation | OLS Coef. |
|---|---|---|
| Risk-taking item | 0.35 | 0.20 |
| Patience item | -0.40 | -0.17 |

Note: Values represent the association between the single-item measures, and behavioural assessments, of risk-taking, and patience utilised in Falk et al., 2018[2]. The Spearman's correlations were calculated using raw data, while the linear regression coefficients were calculated using standardised scores. N = 409. Adapted from Falk et al., 2016[3]. OLS = ordinary least squares.

**Potential confounders**

Confounders included: respondent's sex (male or female); ethnicity (white or non-white); National Statistics Socioeconomic Class prior to the lockdown (NS-SEC, grouped into eight categories: higher managerial, lower managerial, intermediate occupations, small employers and self-employed, lower supervisory and technical, semi-routine occupations, routine occupations, and Uncategorised ), and economic activity during the pandemic (grouped into ten categories: employed, self-employed, unpaid/voluntary work, apprenticeship, unemployed, permanently sick or disabled, looking after the home or family, in education, retired, and Uncategorised ). The Office for National Statistics have published a detailed description of the NS-SEC [4].

The selection of potential confounding variables was driven by the author's substantive knowledge about established risk factors that could plausibly be related to our outcome variables. For instance, there are several sociocultural factors that should be accounted for when researching alcohol misuse using human participants [5]. Historical data suggests that binge drinking is highest among younger individuals and declines with age [6]. However, recently emerging evidence suggests a devaluation of alcohol among Generation Z (born between 1996 and 2015) [7]. Similarly, in Western cultures men tend to drink more than woman, yet data from the US suggests a shift in the pattern, whereby rates of AUD have increased by around 85% among women [8]. One explanation for this may be sex differences in susceptibility to stress [9]. In terms of ethnicity, binge drinking tends to be more prevalent among white people [10]. This is thought to be partly attributable to the way alcohol consumption is often stigmatised among ethnic minorities [11,12]. Nevertheless, due to this stigmatisation, individuals that belong to these cultural groups tend to be disproportionately affected by alcohol-related harm [12]. Further, having a lower socioeconomic status has been previously reported as being associated with lower total alcohol consumption, yet being at the greatest risk of hazardous drinking and alcohol-related harm, perhaps due to higher levels of heavy episodic (binge) drinking among more deprived groups [13,14].

**Missing data**

Weights were derived from logistic regression models by the Centre for Longitudinal Studies team using several variables associated with non-response. For example, sex, ethnicity, social class, cognitive ability, indicators mental health, educational achievement, internet access prior to the web survey, economic activity, indicators of physical health, and non-response during previous sweeps – see Brown et al. [1] for a detailed description of the procedure used to calculate weights.

**Table S3.** Percentage of missing data by variable.

| Variable | Overall | MCS | Next Steps | BCS70 | NCDS |
|---|---|---|---|---|---|
| *n* | 13,453 | 2,644 | 1,852 | 3,997 | 4,960 |
| Sex | 0.00% | 0.00% | 0.00% | 0.00% | 0.00% |
| Ethnicity | 13.29% | 3.82% | 1.84% | 10.56% | 24.82% |
| Relationship status | 2.70% | 3.59% | 2.92% | 2.20% | 2.54% |
| COVID-19 status | 0.01% | 0.00% | 0.00% | 0.03% | 0.02% |
| Economic activity at time of survey | 5.07% | 8.17% | 5.56% | 3.53% | 4.48% |
| Key worker | 5.29% | 8.28% | 5.72% | 0.00% | 4.82% |
| NS–SEC 2010 analytical classes | 0.00% | 0.00% | 0.00% | 0.00% | 0.00% |
| Change in drinking | 7.87% | 9.68% | 6.70% | 6.35% | 8.57% |
| Alcohol misuse at time of survey | 8.55% | 10.25% | 7.67% | 6.96% | 9.25% |
| Change in stress | 8.07% | 12.67% | 9.67% | 6.05% | 6.65% |
| Risk-taking | 7.83% | 12.29% | 8.96% | 3.70% | 6.43% |
| Impatience | 8.02% | 12.41% | 8.96% | 6.08% | 6.88% |
| PHQ–4 | 0.00% | 0.00% | 0.00% | 0.00% | 0.00% |

Note: NS-SEC = National Statistics Socio-economic Class; PHQ–4 = Patient Health Questionnaire – 4. The overall percentage of missing data was 23.43%.

### Supplementary Results

#### Change in alcohol use

**Table S4.** Ordinal logistic regression results for the Millennium Cohort Study with change in alcohol use as the outcome.

| | Model 1 | | | Model 2 | | |
|---|---|---|---|---|---|---|
| **Variable** | **OR (95% CI)** | **SE** | ***p*** | **OR (95% CI)** | **SE** | ***p*** |
| Sex | | | | | | |
| Male | Ref. | | | Ref. | | |
| Female | 0.77 (0.51, 1.14) | 0.16 | 0.190 | 0.83 (0.55, 1.27) | 0.18 | 0.400 |
| Ethnicity | | | | | | |
| White | Ref. | | | Ref. | | |
| Non-white | 1.39 (0.64, 3.01) | 0.55 | 0.404 | 1.58 (0.72, 3.47) | 0.63 | 0.251 |
| NS-SEC analytical classes | | | | | | |
| Higher managerial | Ref. | | | Ref. | | |
| Lower managerial | 3.77 (1.00, 14.23) | 2.54 | 0.050 | 3.27 (0.81, 13.27) | 2.33 | 0.096 |
| Intermediate occupations | 1.81 (0.68, 4.81) | 0.90 | 0.235 | 1.84 (0.67, 5.07) | 0.95 | 0.239 |
| Small employers and self employed | 1.11 (0.27, 4.54) | 0.79 | 0.888 | 1.16 (0.25, 5.31) | 0.90 | 0.851 |
| Lower supervisory and technical | 0.56 (0.18, 1.73) | 0.32 | 0.309 | 0.56 (0.17, 1.85) | 0.34 | 0.336 |
| Semi-routine occupations | 2.13 (0.83, 5.5) | 1.03 | 0.118 | 2.1 (0.77, 5.77) | 1.08 | 0.148 |
| Routine occupations | 1.93 (0.68, 5.43) | 1.01 | 0.213 | 1.62 (0.55, 4.79) | 0.89 | 0.380 |
| Uncategorised | 2.11 (0.72, 6.19) | 1.15 | 0.173 | 2.13 (0.67, 6.8) | 1.26 | 0.201 |
| Economic activity | | | | | | |
| Employed | Ref. | | | Ref. | | |
| Self-employed | 0.56 (0.15, 2.03) | 0.37 | 0.375 | 0.55 (0.15, 2.05) | 0.37 | 0.371 |
| Unpaid/voluntary work | 0.16 (0.02, 1.26) | 0.17 | 0.081 | 0.16 (0.01, 1.74) | 0.19 | 0.131 |
| Apprenticeship | 1.37 (0.71, 2.63) | 0.45 | 0.343 | 1.38 (0.68, 2.79) | 0.49 | 0.370 |
| Unemployed | 0.80 (0.42, 1.54) | 0.27 | 0.505 | 0.70 (0.35, 1.41) | 0.25 | 0.319 |
| Permanently sick or disabled | 0.71 (0.17, 2.96) | 0.51 | 0.637 | 0.54 (0.13, 2.29) | 0.40 | 0.405 |
| Retired | - | - | - | - | - | - |
| Looking after home or family | 0.70 (0.25, 1.97) | 0.37 | 0.504 | 0.48 (0.17, 1.39) | 0.26 | 0.177 |
| In education | **0.10 (0.02, 0.59)** | **0.09** | **0.011** | **0.12 (0.03, 0.49)** | **0.09** | **0.004** |
| Uncategorised | 0.95 (0.52, 1.73) | 0.29 | 0.858 | 0.88 (0.44, 1.75) | 0.31 | 0.709 |
| Stress | | | | | | |
| Same | Ref. | | | Ref. | | |
| Less | | | | 0.21 (0.02, 1.98) | 0.24 | 0.172 |
| More | | | | 1.47 (0.39, 5.61) | 1.00 | 0.568 |
| Risk-taking | | | | 0.98 (0.88, 1.10) | 0.06 | 0.775 |
| Risk-taking x Stress | | | | | | |
| Same | Ref. | | | Ref. | | |
| Less | | | | 1.2 (0.92, 1.57) | 0.16 | 0.181 |
| More | | | | 1.05 (0.87, 1.26) | 0.10 | 0.622 |
| Impatience | | | | **1.14 (1.06, 1.24)** | **0.05** | **0.001** |
| Impatience x Stress | | | | | | |
| Same | Ref. | | | Ref. | | |
| Less | | | | 0.92 (0.75, 1.12) | 0.09 | 0.404 |

| | | | |
|---|---|---|---|
| More | **0.87 (0.77, 0.99)** | **0.06** | **0.030** |

NS-SEC = National Statistics Socio-economic Class. Model 1: Demographics (sex, ethnicity, NS-SEC prior to the outbreak of Coronavirus, and economic activity during the pandemic). Model 2: The effect of inhibitory control (risk-taking and patience), stress, and the interaction between inhibitory control and stress, adjusting for demographics.

**Table S5.** Ordinal logistic regression results for the Next Steps cohort with change in alcohol use as the outcome.

| | Model 1 | | | Model 2 | | |
|---|---|---|---|---|---|---|
| **Variable** | **OR (95% CI)** | **SE** | ***p*** | **OR (95% CI)** | **SE** | ***p*** |
| Sex | | | | | | |
| Male | Ref. | | | Ref. | | |
| Female | 1.14 (0.85, 1.54) | 0.17 | 0.380 | 1.17 (0.86, 1.59) | 0.18 | 0.315 |
| Ethnicity | | | | | | |
| White | Ref. | | | Ref. | | |
| Non-white | **0.71 (0.55, 0.93)** | **0.10** | **0.012** | **0.70 (0.54, 0.91)** | **0.09** | **0.008** |
| NS-SEC analytical classes | | | | | | |
| Higher managerial | Ref. | | | Ref. | | |
| Lower managerial | 0.90 (0.61, 1.32) | 0.18 | 0.575 | 0.91 (0.62, 1.34) | 0.18 | 0.640 |
| Intermediate occupations | 0.72 (0.45, 1.14) | 0.17 | 0.161 | 0.64 (0.39, 1.05) | 0.16 | 0.075 |
| Small employers and self employed | 0.96 (0.39, 2.39) | 0.44 | 0.937 | 0.88 (0.35, 2.23) | 0.42 | 0.788 |
| Lower supervisory and technical | 0.90 (0.31, 2.6) | 0.49 | 0.850 | 0.91 (0.33, 2.5) | 0.47 | 0.851 |
| Semi-routine occupations | 1.11 (0.63, 1.97) | 0.32 | 0.713 | 1.19 (0.65, 2.15) | 0.36 | 0.574 |
| Routine occupations | 1.01 (0.52, 1.96) | 0.34 | 0.985 | 1.14 (0.51, 2.54) | 0.46 | 0.743 |
| Uncategorised | 1.59 (0.81, 3.1) | 0.54 | 0.175 | 1.36 (0.68, 2.7) | 0.48 | 0.384 |
| Economic activity | | | | | | |
| Employed | Ref. | | | Ref. | | |
| Self-employed | 1.36 (0.82, 2.25) | 0.35 | 0.235 | 1.22 (0.73, 2.04) | 0.32 | 0.458 |
| Unpaid/voluntary work | 0.39 (0.05, 2.99) | 0.40 | 0.363 | 0.47 (0.06, 3.62) | 0.49 | 0.467 |
| Apprenticeship | 0.79 (0.54, 1.15) | 0.15 | 0.212 | 0.89 (0.59, 1.33) | 0.18 | 0.562 |
| Unemployed | 0.45 (0.19, 1.06) | 0.20 | 0.069 | 0.51 (0.2, 1.29) | 0.24 | 0.155 |
| Permanently sick or disabled | 0.39 (0.11, 1.35) | 0.25 | 0.138 | 0.44 (0.12, 1.57) | 0.29 | 0.207 |
| Retired | - | - | - | - | - | - |
| Looking after home or family | 0.47 (0.14, 1.54) | 0.28 | 0.213 | 0.58 (0.17, 1.94) | 0.36 | 0.377 |
| In education | - | - | - | - | - | - |
| Uncategorised | **0.42 (0.17, 0.99)** | **0.19** | **0.049** | 0.45 (0.18, 1.11) | 0.21 | 0.083 |
| Stress | | | | | | |
| Same | Ref. | | | Ref. | | |
| Less | | | | 0.59 (0.09, 3.71) | 0.55 | 0.574 |
| More | | | | 2.21 (0.99, 4.94) | 0.90 | 0.053 |
| Risk-taking | | | | 1.03 (0.95, 1.13) | 0.05 | 0.479 |
| Risk-taking x Stress | | | | | | |
| Same | Ref. | | | Ref. | | |
| Less | | | | 0.96 (0.74, 1.24) | 0.13 | 0.760 |
| More | | | | 0.98 (0.87, 1.09) | 0.06 | 0.676 |
| Impatience | | | | 1.05 (0.97, 1.14) | 0.04 | 0.201 |
| Impatience x Stress | | | | | | |
| Same | Ref. | | | Ref. | | |
| Less | | | | **1.22 (1.00, 1.48)** | **0.12** | **0.047** |
| More | | | | **0.88 (0.80, 0.98)** | **0.05** | **0.016** |

NS-SEC = National Statistics Socio-economic Class. Model 1: Demographics (sex, ethnicity, NS-SEC prior to the outbreak of Coronavirus, and economic activity during the pandemic). Model 2: The effect of inhibitory control (risk-taking and patience), stress, and the interaction between inhibitory control and stress, adjusting for demographics.

**Table S6.** Ordinal logistic regression results for the 1970 British Cohort Study with change in alcohol use as the outcome.

| | **Model 1** | | | **Model 2** | | |
|---|---|---|---|---|---|---|
| **Variable** | **OR (95% CI)** | **SE** | ***p*** | **OR (95% CI)** | **SE** | ***p*** |
| Sex | | | | | | |
| Male | Ref. | | | Ref. | | |
| Female | **1.27 (1.08, 1.50)** | **0.11** | **0.004** | **1.19 (1.01, 1.41)** | **0.10** | **0.043** |
| Ethnicity | | | | | | |
| White | Ref. | | | Ref. | | |
| Non-white | 0.77 (0.51, 1.16) | 0.16 | 0.205 | 0.78 (0.51, 1.18) | 0.16 | 0.234 |
| NS-SEC analytical classes | | | | | | |
| Higher managerial | Ref. | | | Ref. | | |
| Lower managerial | 0.96 (0.76, 1.22) | 0.12 | 0.753 | 0.95 (0.75, 1.22) | 0.12 | 0.708 |
| Intermediate occupations | **0.70 (0.54, 0.92)** | **0.10** | **0.010** | **0.70 (0.53, 0.92)** | **0.10** | **0.010** |
| Small employers and self employed | 0.78 (0.49, 1.23) | 0.18 | 0.287 | 0.83 (0.52, 1.34) | 0.20 | 0.444 |
| Lower supervisory and technical | 0.99 (0.65, 1.51) | 0.21 | 0.955 | 1.02 (0.66, 1.58) | 0.23 | 0.932 |
| Semi-routine occupations | **0.62 (0.46, 0.85)** | **0.10** | **0.003** | **0.59 (0.43, 0.81)** | **0.10** | **0.001** |
| Routine occupations | **0.62 (0.39, 0.98)** | **0.15** | **0.041** | **0.56 (0.36, 0.87)** | **0.12** | **0.009** |
| Uncategorised | 1.02 (0.68, 1.54) | 0.21 | 0.912 | 1.17 (0.8, 1.71) | 0.23 | 0.432 |
| Economic activity | | | | | | |
| Employed | Ref. | | | Ref. | | |
| Self-employed | 1.11 (0.85, 1.45) | 0.15 | 0.435 | 0.98 (0.76, 1.25) | 0.12 | 0.851 |
| Unpaid/voluntary work | 1.44 (0.33, 6.35) | 1.09 | 0.633 | 1.37 (0.32, 5.94) | 1.03 | 0.673 |
| Apprenticeship | - | - | - | - | - | - |
| Unemployed | 0.73 (0.43, 1.25) | 0.20 | 0.253 | 0.66 (0.38, 1.15) | 0.19 | 0.145 |
| Permanently sick or disabled | **0.40 (0.24, 0.66)** | **0.10** | **< 0.001** | **0.35 (0.21, 0.56)** | **0.09** | **< 0.001** |
| Retired | 1.09 (0.67, 1.75) | 0.27 | 0.732 | 0.97 (0.61, 1.55) | 0.23 | 0.900 |
| Looking after home or family | 0.87 (0.47, 1.6) | 0.27 | 0.655 | 0.91 (0.49, 1.7) | 0.29 | 0.766 |
| In education | **-** | **-** | **-** | - | - | - |
| Uncategorised | **0.36 (0.14, 0.94)** | **0.18** | **0.036** | **0.33 (0.12, 0.9)** | **0.17** | **0.031** |
| Stress | | | | | | |
| Same | Ref. | | | Ref. | | |
| Less | | | | 1.40 (0.58, 3.38) | 0.63 | 0.455 |
| More | | | | 0.87 (0.51, 1.47) | 0.23 | 0.594 |
| Risk-taking | | | | 0.98 (0.94, 1.03) | 0.02 | 0.533 |
| Risk-taking x Stress | | | | | | |
| Same | Ref. | | | Ref. | | |
| Less | | | | 0.96 (0.85, 1.08) | 0.06 | 0.478 |
| More | | | | 1.06 (0.98, 1.15) | 0.04 | 0.152 |
| Impatience | | | | 0.98 (0.93, 1.03) | 0.02 | 0.370 |
| Impatience x Stress | | | | | | |
| Same | Ref. | | | Ref. | | |
| Less | | | | 1.01 (0.9, 1.14) | 0.06 | 0.846 |
| More | | | | 1.05 (0.97, 1.13) | 0.04 | 0.216 |

NS-SEC = National Statistics Socio-economic Class. Model 1: Demographics (sex, ethnicity, NS-SEC prior to the outbreak of Coronavirus, and economic activity during the pandemic). Model 2: The effect of inhibitory control (risk-taking and patience), stress, and the interaction between inhibitory control and stress, adjusting for demographics.

**Table S7.** Ordinal logistic regression results for the National Child Development Study with change in alcohol use as the outcome.

| | Model 1 | | | Model 2 | | |
|---|---|---|---|---|---|---|
| **Variable** | **OR (95% CI)** | **SE** | ***p*** | **OR (95% CI)** | **SE** | ***p*** |
| Sex | | | | | | |
| Male | Ref. | | | Ref. | | |
| Female | **1.23 (1.02, 1.50)** | **0.12** | **0.035** | 1.19 (0.98, 1.44) | 0.12 | 0.081 |
| Ethnicity | | | | | | |
| White | Ref. | | | Ref. | | |
| Non-white | 0.54 (0.26, 1.09) | 0.19 | 0.083 | 0.53 (0.26, 1.08) | 0.19 | 0.081 |
| NS-SEC analytical classes | | | | | | |
| Higher managerial | Ref. | | | Ref. | | |
| Lower managerial | 0.85 (0.6, 1.2) | 0.15 | 0.354 | 0.88 (0.61, 1.25) | 0.16 | 0.467 |
| Intermediate occupations | 0.9 (0.62, 1.31) | 0.17 | 0.593 | 0.91 (0.62, 1.32) | 0.17 | 0.610 |
| Small employers and self employed | 0.73 (0.44, 1.22) | 0.19 | 0.235 | 0.81 (0.5, 1.34) | 0.21 | 0.418 |
| Lower supervisory and technical | **0.45 (0.24, 0.84)** | **0.14** | **0.012** | **0.45 (0.24, 0.86)** | **0.15** | **0.015** |
| Semi-routine occupations | 0.96 (0.59, 1.57) | 0.24 | 0.870 | 1.03 (0.63, 1.69) | 0.26 | 0.896 |
| Routine occupations | 0.76 (0.48, 1.22) | 0.18 | 0.260 | 0.79 (0.49, 1.28) | 0.19 | 0.334 |
| Uncategorised | 0.7 (0.4, 1.22) | 0.20 | 0.206 | 0.71 (0.39, 1.28) | 0.21 | 0.257 |
| Economic activity | | | | | | |
| Employed | Ref. | | | Ref. | | |
| Self-employed | 0.94 (0.7, 1.26) | 0.14 | 0.669 | 0.91 (0.66, 1.25) | 0.15 | 0.566 |
| Unpaid/voluntary work | 1.61 (0.45, 5.71) | 1.04 | 0.462 | 1.51 (0.42, 5.46) | 0.99 | 0.529 |
| Apprenticeship | - | - | - | - | - | - |
| Unemployed | 0.69 (0.21, 2.22) | 0.41 | 0.532 | 0.65 (0.21, 2) | 0.37 | 0.453 |
| Permanently sick or disabled | 1.11 (0.61, 2.02) | 0.34 | 0.743 | 1.13 (0.6, 2.13) | 0.37 | 0.706 |
| Retired | 1.00 (0.59, 1.71) | 0.27 | 0.988 | 1.01 (0.58, 1.75) | 0.28 | 0.978 |
| Looking after home or family | 1.04 (0.55, 1.98) | 0.34 | 0.903 | 1.05 (0.54, 2.06) | 0.36 | 0.881 |
| In education | - | - | - | - | - | - |
| Uncategorised | 1.08 (0.32, 3.7) | 0.68 | 0.902 | 1.04 (0.28, 3.91) | 0.70 | 0.957 |
| Stress | | | | | | |
| Same | Ref. | | | Ref. | | |
| Less | | | | 1.38 (0.42, 4.51) | 0.83 | 0.590 |
| More | | | | 0.90 (0.54, 1.48) | 0.23 | 0.670 |
| Risk-taking | | | | 0.99 (0.94, 1.03) | 0.02 | 0.508 |
| Risk-taking x Stress | | | | | | |
| Same | Ref. | | | Ref. | | |
| Less | | | | 0.97 (0.82, 1.13) | 0.08 | 0.674 |
| More | | | | 1.07 (0.98, 1.16) | 0.04 | 0.118 |
| Impatience | | | | 0.99 (0.95, 1.03) | 0.02 | 0.504 |
| Impatience x Stress | | | | | | |
| Same | Ref. | | | Ref. | | |
| Less | | | | 1.01 (0.89, 1.15) | 0.07 | 0.869 |
| More | | | | 0.99 (0.93, 1.07) | 0.04 | 0.875 |

NS-SEC = National Statistics Socio-economic Class. Model 1: Demographics (sex, ethnicity, NS-SEC prior to the outbreak of Coronavirus, and economic activity during the pandemic). Model 2: The effect of inhibitory control (risk-taking and patience), stress, and the interaction between inhibitory control and stress, adjusting for demographics.

**Risk of alcohol-related harm due to hazardous drinking**

**Table S8.** Ordinal logistic regression results for the Millennium Cohort Study with risk of alcohol-related harm due to hazardous drinking as the outcome.

| | **Model 1** | | | **Model 2** | | |
|---|---|---|---|---|---|---|
| **Variable** | **OR (95% CI)** | **SE** | ***p*** | **OR (95% CI)** | **SE** | ***p*** |
| Sex | | | | | | |
| Male | Ref. | | | Ref. | | |
| Female | 0.60 (0.36, 1.02) | 0.16 | 0.062 | 0.58 (0.34, 1.02) | 0.16 | 0.057 |
| Ethnicity | | | | | | |
| White | Ref. | | | Ref. | | |
| Non-white | 0.55 (0.23, 1.33) | 0.25 | 0.185 | 0.71 (0.3, 1.67) | 0.31 | 0.427 |
| NS-SEC analytical classes | | | | | | |
| Higher managerial | Ref. | | | Ref. | | |
| Lower managerial | 1.76 (0.39, 8.04) | 1.36 | 0.463 | 1.47 (0.28, 7.77) | 1.24 | 0.649 |
| Intermediate occupations | 0.67 (0.17, 2.59) | 0.46 | 0.557 | 0.71 (0.16, 3.15) | 0.54 | 0.655 |
| Small employers and self employed | 1.46 (0.33, 6.5) | 1.11 | 0.618 | 1.04 (0.21, 5.14) | 0.84 | 0.963 |
| Lower supervisory and technical | 0.34 (0.08, 1.53) | 0.26 | 0.159 | 0.29 (0.06, 1.48) | 0.24 | 0.135 |
| Semi-routine occupations | 0.74 (0.18, 3.09) | 0.54 | 0.681 | 0.69 (0.14, 3.36) | 0.56 | 0.647 |
| Routine occupations | 0.68 (0.17, 2.75) | 0.48 | 0.588 | 0.56 (0.12, 2.71) | 0.45 | 0.472 |
| Uncategorised | 1.08 (0.27, 4.33) | 0.76 | 0.917 | 1.36 (0.3, 6.21) | 1.05 | 0.687 |
| Economic activity | | | | | | |
| Employed | Ref. | | | Ref. | | |
| Self-employed | 0.84 (0.27, 2.58) | 0.48 | 0.757 | 0.65 (0.2, 2.17) | 0.40 | 0.485 |
| Unpaid/voluntary work | **5.60E-07 (1.30E-07, 2.42E-06)** | **4.16E-07** | **< 0.001** | **6.40E-07 (1.04E-07, 3.96E-06)** | **5.92E-07** | **< 0.001** |
| Apprenticeship | **0.29 (0.11, 0.78)** | **0.15** | **0.015** | **0.25 (0.08, 0.77)** | **0.14** | **0.016** |
| Unemployed | 0.79 (0.39, 1.61) | 0.29 | 0.521 | 0.72 (0.37, 1.4) | 0.24 | 0.333 |
| Permanently sick or disabled | **0.11 (0.01, 1.03)** | **0.12** | **0.053** | **0.07 (0.01, 0.68)** | **0.08** | **0.021** |
| Retired | - | - | - | - | - | - |
| Looking after home or family | **6.91E-07 (2.37E-07, 2.01E-06)** | **3.75E-07,** | **< 0.001** | **5.86E-07 (9.92E-08, 3.46E-06)** | **5.29E-07** | **< 0.001** |
| In education | **2.63 (1.37, 5.05)** | **0.87** | **0.004** | 2.16 (0.93, 5.04) | 0.93 | 0.075 |
| Uncategorised | **0.18 (0.05, 0.62)** | **0.11** | **0.007** | **0.11 (0.03, 0.46)** | **0.08** | **0.003** |
| Stress | | | | | | |
| Same | Ref. | | | Ref. | | |
| Less | | | | 0.25 (0.01, 4.98) | 0.38 | 0.361 |
| More | | | | 0.82 (0.18, 3.65) | 0.62 | 0.794 |
| Risk-taking | | | | 0.98 (0.8, 1.19) | 0.10 | 0.836 |
| Risk-taking x Stress | | | | | | |
| Same | Ref. | | | Ref. | | |
| Less | | | | 1.32 (0.89, 1.96) | 0.27 | 0.172 |
| More | | | | 1.13 (0.91, 1.41) | 0.12 | 0.258 |
| Impatience | | | | **1.20 (1.05, 1.38)** | **0.08** | **0.010** |
| Impatience x Stress | | | | | | |
| Same | Ref. | | | Ref. | | |
| Less | | | | 0.9 (0.73, 1.12) | 0.10 | 0.359 |

| | | | |
|---|---|---|---|
| More | 0.95 (0.8, 1.14) | 0.09 | 0.603 |

NS-SEC = National Statistics Socio-economic Class. Model 1: Demographics (sex, ethnicity, NS-SEC prior to the outbreak of Coronavirus, and economic activity during the pandemic). Model 2: The effect of inhibitory control (risk-taking and patience), stress, and the interaction between inhibitory control and stress, adjusting for demographics.

**Table S9.** Ordinal logistic regression results for the Next Steps cohort with risk of alcohol-related harm due to hazardous drinking as the outcome.

| | Model 1 | | | Model 2 | | |
|---|---|---|---|---|---|---|
| **Variable** | **OR (95% CI)** | **SE** | ***p*** | **OR (95% CI)** | **SE** | ***p*** |
| Sex | | | | | | |
| Male | Ref. | | | Ref. | | |
| Female | **0.60 (0.42, 0.85)** | **0.11** | **0.004** | **0.56 (0.39, 0.82)** | **0.11** | **0.003** |
| Ethnicity | | | | | | |
| White | Ref. | | | Ref. | | |
| Non-white | **0.55 (0.34, 0.91)** | **0.14** | **0.02** | **0.44 (0.27, 0.73)** | **0.11** | **0.002** |
| NS-SEC analytical classes | | | | | | |
| Higher managerial | Ref. | | | Ref. | | |
| Lower managerial | 1.03 (0.65, 1.63) | 0.24 | 0.893 | 1.02 (0.65, 1.6) | 0.24 | 0.935 |
| Intermediate occupations | 0.6 (0.37, 0.97) | 0.15 | 0.037 | 0.59 (0.35, 0.99) | 0.16 | 0.047 |
| Small employers and self employed | 1.43 (0.57, 3.58) | 0.67 | 0.448 | 1.2 (0.41, 3.52) | 0.66 | 0.741 |
| Lower supervisory and technical | 1.03 (0.36, 2.93) | 0.55 | 0.956 | 1.00 (0.35, 2.87) | 0.54 | 0.998 |
| Semi-routine occupations | 0.99 (0.49, 1.97) | 0.35 | 0.971 | 0.93 (0.44, 1.94) | 0.35 | 0.845 |
| Routine occupations | 0.55 (0.23, 1.32) | 0.25 | 0.181 | 0.62 (0.24, 1.61) | 0.3 | 0.327 |
| Uncategorised | 0.63 (0.23, 1.7) | 0.32 | 0.357 | 0.6 (0.21, 1.69) | 0.32 | 0.328 |
| Economic activity | | | | | | |
| Employed | Ref. | | | Ref. | | |
| Self-employed | 1.19 (0.64, 2.22) | 0.38 | 0.584 | 1.12 (0.57, 2.2) | 0.38 | 0.732 |
| Unpaid/voluntary work | 0.66 (0.03, 14.5) | 1.04 | 0.793 | 0.64 (0.02, 19.07) | 1.1 | 0.795 |
| Apprenticeship | 1.73 (0.41, 7.28) | 1.27 | 0.454 | 2.53 (0.62, 10.32) | 1.81 | 0.196 |
| Unemployed | 1.78 (0.56, 5.68) | 1.05 | 0.327 | 2.20 (0.72, 6.74) | 1.25 | 0.167 |
| Permanently sick or disabled | 0.68 (0.08, 5.73) | 0.74 | 0.721 | 0.91 (0.11, 7.71) | 0.99 | 0.928 |
| Retired | - | - | - | - | - | - |
| Looking after home or family | 1.73 (0.36, 8.33) | 1.38 | 0.496 | 2.38 (0.44, 12.94) | 2.05 | 0.315 |
| In education | - | - | - | - | - | - |
| Uncategorised | 0.64 (0.13, 3.24) | 0.53 | 0.589 | 0.66 (0.13, 3.42) | 0.55 | 0.617 |
| Stress | | | | | | |
| Same | Ref. | | | Ref. | | |
| Less | | | | 0.36 (0.06, 2.15) | 0.33 | 0.259 |
| More | | | | **3.77 (1.15, 12.28)** | **2.27** | **0.028** |
| Risk-taking | | | | **1.18 (1.05, 1.32)** | **0.07** | **0.006** |
| Risk-taking x Stress | | | | | | |
| Same | Ref. | | | Ref. | | |
| Less | | | | 0.97 (0.77, 1.22) | 0.11 | 0.813 |
| More | | | | 0.88 (0.77, 1.02) | 0.07 | 0.098 |
| Impatience | | | | 0.97 (0.9, 1.06) | 0.04 | 0.531 |
| Impatience x Stress | | | | | | |
| Same | Ref. | | | Ref. | | |
| Less | | | | **1.31 (1.10, 1.57)** | **0.12** | **0.002** |
| More | | | | 0.95 (0.83, 1.07) | 0.06 | 0.396 |

NS-SEC = National Statistics Socio-economic Class. Model 1: Demographics (sex, ethnicity, NS-SEC prior to the outbreak of Coronavirus, and economic activity during the pandemic). Model 2: The effect of inhibitory control (risk-taking and patience), stress, and the interaction between inhibitory control and stress, adjusting for demographics.

**Table S10.** Ordinal logistic regression results for the 1970 British Cohort Study with risk of alcohol-related harm due to hazardous drinking as the outcome.

| | **Model 1** | | | **Model 2** | | |
|---|---|---|---|---|---|---|
| **Variable** | **OR (95% CI)** | **SE** | ***p*** | **OR (95% CI)** | **SE** | ***p*** |
| Sex | | | | | | |
| Male | Ref. | | | Ref. | | |
| Female | **0.64 (0.53, 0.76)** | **0.06** | **< 0.001** | **0.64 (0.53, 0.77)** | **0.06** | **< 0.001** |
| Ethnicity | | | | | | |
| White | Ref. | | | Ref. | | |
| Non-white | **0.44 (0.23, 0.84)** | **0.15** | **< 0.001** | **0.41 (0.21, 0.81)** | **0.14** | **0.010** |
| NS-SEC analytical classes | | | | | | |
| Higher managerial | Ref. | | | Ref. | | |
| Lower managerial | 1.03 (0.83, 1.29) | 0.12 | 0.775 | 1.03 (0.82, 1.29) | 0.12 | 0.808 |
| Intermediate occupations | 0.79 (0.6, 1.04) | 0.11 | 0.098 | 0.82 (0.62, 1.08) | 0.12 | 0.164 |
| Small employers and self employed | 0.85 (0.55, 1.32) | 0.19 | 0.478 | 0.87 (0.56, 1.36) | 0.20 | 0.536 |
| Lower supervisory and technical | 0.70 (0.42, 1.18) | 0.19 | 0.183 | 0.76 (0.45, 1.27) | 0.20 | 0.289 |
| Semi-routine occupations | 0.85 (0.61, 1.18) | 0.14 | 0.335 | 0.84 (0.6, 1.18) | 0.15 | 0.321 |
| Routine occupations | 0.70 (0.44, 1.11) | 0.17 | 0.132 | **0.65 (0.44, 0.96)** | **0.13** | **0.030** |
| Uncategorised | 0.91 (0.6, 1.38) | 0.19 | 0.652 | 1.02 (0.68, 1.53) | 0.21 | 0.939 |
| Economic activity | | | | | | |
| Employed | Ref. | | | Ref. | | |
| Self-employed | 1.1 (0.82, 1.48) | 0.16 | 0.512 | 1.01 (0.77, 1.32) | 0.14 | 0.944 |
| Unpaid/voluntary work | 1.12 (0.24, 5.14) | 0.87 | 0.884 | 1.12 (0.24, 5.1) | 0.87 | 0.886 |
| Apprenticeship | - | - | - | - | - | - |
| Unemployed | 0.83 (0.43, 1.6) | 0.28 | 0.584 | 0.77 (0.41, 1.45) | 0.25 | 0.422 |
| Permanently sick or disabled | **0.24 (0.10, 0.58)** | **0.11** | **0.002** | **0.21 (0.09, 0.53)** | **0.10** | **0.001** |
| Retired | 0.89 (0.54, 1.47) | 0.23 | 0.653 | 0.82 (0.49, 1.36) | 0.21 | 0.437 |
| Looking after home or family | 0.7 (0.26, 1.86) | 0.35 | 0.475 | 0.72 (0.28, 1.85) | 0.35 | 0.498 |
| In education | - | - | - | - | - | - |
| Uncategorised | 0.7 (0.28, 1.75) | 0.33 | 0.442 | 0.69 (0.27, 1.78) | 0.33 | 0.442 |
| Stress | | | | | | |
| Same | Ref. | | | Ref. | | |
| Less | | | | 1.00 (0.35, 2.89) | 0.54 | 0.999 |
| More | | | | 1.29 (0.73, 2.25) | 0.37 | 0.380 |
| Risk-taking | | | | **1.06 (1.01, 1.12)** | **0.03** | **0.017** |
| Risk-taking x Stress | | | | | | |
| Same | Ref. | | | Ref. | | |
| Less | | | | 0.95 (0.82, 1.1) | 0.07 | 0.504 |
| More | | | | 1.01 (0.93, 1.09) | 0.04 | 0.834 |
| Impatience | | | | 1.00 (0.95, 1.04) | 0.02 | 0.859 |
| Impatience x Stress | | | | | | |
| Same | Ref. | | | Ref. | | |
| Less | | | | **1.17 (1.04, 1.31)** | **0.07** | **0.007** |
| More | | | | 1.00 (0.93, 1.08) | 0.04 | 0.943 |

NS-SEC = National Statistics Socio-economic Class. Model 1: Demographics (sex, ethnicity, NS-SEC prior to the outbreak of Coronavirus, and economic activity during the pandemic). Model 2: The effect of inhibitory control (risk-taking and patience), stress, and the interaction between inhibitory control and stress, adjusting for demographics.

**Table S11.** Ordinal logistic regression results for the National Child Development Study with risk of alcohol-related harm due to hazardous drinking as the outcome.

| | Model 1 | | | Model 2 | | |
|---|---|---|---|---|---|---|
| **Variable** | **OR (95% CI)** | **SE** | ***p*** | **OR (95% CI)** | **SE** | ***p*** |
| Sex | | | | | | |
| Male | Ref. | | | Ref. | | |
| Female | **0.64 (0.52, 0.78)** | **0.07** | **< 0.001** | **0.62 (0.5, 0.76)** | **0.07** | **< 0.001** |
| Ethnicity | | | | | | |
| White | Ref. | | | Ref. | | |
| Non-white | **0.26 (0.12, 0.56)** | **0.10** | **< 0.001** | **0.27 (0.13, 0.58)** | **0.11** | **< 0.001** |
| NS-SEC analytical classes | | | | | | |
| Higher managerial | Ref. | | | Ref. | | |
| Lower managerial | 0.84 (0.57, 1.24) | 0.17 | 0.378 | 0.82 (0.56, 1.19) | 0.16 | 0.295 |
| Intermediate occupations | 0.75 (0.51, 1.12) | 0.15 | 0.160 | 0.75 (0.51, 1.1) | 0.15 | 0.138 |
| Small employers and self employed | 0.71 (0.39, 1.28) | 0.21 | 0.256 | 0.75 (0.42, 1.33) | 0.22 | 0.324 |
| Lower supervisory and technical | 0.66 (0.38, 1.13) | 0.18 | 0.133 | 0.71 (0.42, 1.22) | 0.19 | 0.213 |
| Semi-routine occupations | 0.83 (0.49, 1.4) | 0.22 | 0.483 | 0.83 (0.49, 1.41) | 0.22 | 0.500 |
| Routine occupations | **0.56 (0.33, 0.96)** | **0.15** | **0.035** | **0.55 (0.32, 0.96)** | **0.16** | **0.035** |
| Uncategorised | 0.85 (0.52, 1.37) | 0.21 | 0.501 | 0.93 (0.57, 1.52) | 0.23 | 0.784 |
| Economic activity | | | | | | |
| Employed | Ref. | | | Ref. | | |
| Self-employed | 0.74 (0.51, 1.08) | 0.14 | 0.120 | 0.66 (0.46, 0.96) | 0.12 | 0.029 |
| Unpaid/voluntary work | 1.36 (0.67, 2.78) | 0.50 | 0.394 | 1.32 (0.64, 2.73) | 0.49 | 0.449 |
| Apprenticeship | - | - | - | - | - | - |
| Unemployed | 0.87 (0.37, 2.08) | 0.39 | 0.761 | 0.77 (0.32, 1.82) | 0.34 | 0.548 |
| Permanently sick or disabled | 0.59 (0.26, 1.33) | 0.24 | 0.203 | 0.45 (0.2, 1.01) | 0.19 | 0.054 |
| Retired | 1.05 (0.7, 1.59) | 0.22 | 0.807 | 0.88 (0.4, 1.93) | 0.35 | 0.750 |
| Looking after home or family | 0.76 (0.44, 1.3) | 0.21 | 0.312 | 0.95 (0.62, 1.44) | 0.20 | 0.797 |
| In education | - | - | - | - | - | - |
| Uncategorised | 0.92 (0.42, 2) | 0.36 | 0.831 | 0.88 (0.4, 1.93) | 0.35 | 0.750 |
| Stress | | | | | | |
| Same | Ref. | | | Ref. | | |
| Less | | | | 0.74 (0.25, 2.16) | 0.41 | 0.585 |
| More | | | | 0.88 (0.49, 1.60) | 0.27 | 0.680 |
| Risk-taking | | | | 1.00 (0.95, 1.05) | 0.03 | 0.945 |
| Risk-taking x Stress | | | | | | |
| Same | Ref. | | | Ref. | | |
| Less | | | | 1.04 (0.89, 1.22) | 0.08 | 0.631 |
| More | | | | 1.08 (0.99, 1.18) | 0.05 | 0.091 |
| Impatience | | | | 1.02 (0.97, 1.06) | 0.02 | 0.480 |
| Impatience x Stress | | | | | | |
| Same | Ref. | | | Ref. | | |
| Less | | | | 1.04 (0.94, 1.16) | 0.06 | 0.435 |
| More | | | | 1.00 (0.92, 1.09) | 0.04 | 0.972 |

NS-SEC = National Statistics Socio-economic Class. Model 1: Demographics (sex, ethnicity, NS-SEC prior to the outbreak of Coronavirus, and economic activity during the pandemic). Model 2: The effect of inhibitory control (risk-taking and patience), stress, and the interaction between inhibitory control and stress, adjusting for demographics.

#### Change in stress

**Table S12.** Ordinal logistic regression results for the Millennium Cohort Study with change in stress as the outcome.

| Variable | OR (95% CI) | SE | *p* |
|---|---|---|---|
| Sex | | | |
| Male | Ref. | | |
| Female | **1.54 (1.08, 2.20)** | **0.28** | **0.017** |
| Ethnicity | | | |
| White | Ref. | | |
| Non-white | 1.66 (0.75, 3.66) | 0.67 | 0.213 |
| NS-SEC analytical classes | | | |
| Higher managerial | Ref. | | |
| Lower managerial | 1.11 (0.26, 4.8) | 0.83 | 0.885 |
| Intermediate occupations | 0.7 (0.17, 2.85) | 0.5 | 0.613 |
| Small employers and self employed | 0.81 (0.13, 5.1) | 0.76 | 0.818 |
| Lower supervisory and technical | 0.32 (0.07, 1.39) | 0.24 | 0.128 |
| Semi-routine occupations | 0.79 (0.19, 3.29) | 0.57 | 0.749 |
| Routine occupations | 0.55 (0.12, 2.45) | 0.42 | 0.431 |
| Uncategorised | 0.39 (0.09, 1.67) | 0.29 | 0.205 |
| Economic activity | | | |
| Employed | Ref. | | |
| Self-employed | **5.53 (1.56, 19.57)** | **3.55** | **0.008** |
| Unpaid/voluntary work | 5.33 (0.27, 104.22) | 8.05 | 0.269 |
| Apprenticeship | 0.54 (0.28, 1.02) | 0.17 | 0.056 |
| Unemployed | **1.75 (1.08, 2.83)** | **0.43** | **0.024** |
| Permanently sick or disabled | 1.36 (0.47, 3.92) | 0.73 | 0.567 |
| Retired | - | - | - |
| Looking after home or family | 0.99 (0.35, 2.8) | 0.52 | 0.979 |
| In education | 0.39 (0.04, 3.61) | 0.44 | 0.407 |
| Uncategorised | 0.85 (0.46, 1.59) | 0.27 | 0.618 |

NS-SEC = National Statistics Socio-economic Class.

**Table S13.** Ordinal logistic regression results for the Next Steps cohort with change in stress as the outcome.

| **Variable** | **OR (95% CI)** | **SE** | ***p*** |
|---|---|---|---|
| Sex | | | |
| Male | Ref. | | |
| Female | **1.93 (1.39, 2.70)** | **0.33** | **< 0.001** |
| Ethnicity | | | |
| White | Ref. | | |
| Non-white | 0.93 (0.66, 1.32) | 0.17 | 0.691 |
| NS-SEC analytical classes | | | |
| Higher managerial | Ref. | | |
| Lower managerial | 0.80 (0.52, 1.23) | 0.18 | 0.314 |
| Intermediate occupations | 1.26 (0.77, 2.08) | 0.32 | 0.355 |
| Small employers and self employed | 0.48 (0.18, 1.29) | 0.24 | 0.148 |
| Lower supervisory and technical | 0.61 (0.36, 1.02) | 0.16 | 0.060 |
| Semi-routine occupations | 1.30 (0.79, 2.15) | 0.33 | 0.308 |
| Routine occupations | 1.76 (0.87, 3.56) | 0.63 | 0.114 |
| Uncategorised | 1.35 (0.68, 2.64) | 0.46 | 0.388 |
| Economic activity | | | |
| Employed | Ref. | | |
| Self-employed | **2.14 (1.15, 3.98)** | **0.68** | **0.017** |
| Unpaid/voluntary work | 2.77 (0.4, 19.05) | 2.72 | 0.300 |
| Apprenticeship | 0.36 (0.04, 3.42) | 0.41 | 0.375 |
| Unemployed | 1.26 (0.55, 2.91) | 0.54 | 0.586 |
| Permanently sick or disabled | 0.57 (0.2, 1.66) | 0.31 | 0.306 |
| Retired | - | - | - |
| Looking after home or family | 0.71 (0.29, 1.73) | 0.32 | 0.452 |
| In education | - | - | - |
| Uncategorised | 0.87 (0.39, 1.96) | 0.36 | 0.734 |

NS-SEC = National Statistics Socio-economic Class.

**Table S14.** Ordinal logistic regression results for the 1970 British Cohort Study with change in stress as the outcome.

| **Variable** | **OR (95% CI)** | **SE** | ***p*** |
|---|---|---|---|
| Sex | | | |
| Male | Ref. | | |
| Female | **1.62 (1.37, 1.92)** | **0.14** | **< 0.001** |
| Ethnicity | | | |
| White | Ref. | | |
| Non-white | 0.72 (0.4, 1.3) | 0.22 | 0.280 |
| NS-SEC analytical classes | | | |
| Higher managerial | Ref. | | |
| Lower managerial | 1.08 (0.86, 1.36) | 0.13 | 0.498 |
| Intermediate occupations | 1.13 (0.88, 1.44) | 0.14 | 0.335 |
| Small employers and self employed | 1.11 (0.74, 1.66) | 0.23 | 0.607 |
| Lower supervisory and technical | 0.89 (0.58, 1.37) | 0.19 | 0.602 |
| Semi-routine occupations | 1.28 (0.92, 1.78) | 0.22 | 0.151 |
| Routine occupations | 0.92 (0.69, 1.22) | 0.13 | 0.549 |
| Uncategorised | 1.08 (0.71, 1.63) | 0.23 | 0.728 |
| Economic activity | | | |
| Employed | Ref. | | |
| Self-employed | 1.21 (0.92, 1.6) | 0.17 | 0.180 |
| Unpaid/voluntary work | 1.58 (0.36, 6.8) | 1.18 | 0.542 |
| Apprenticeship | - | - | - |
| Unemployed | 1.33 (0.63, 2.8) | 0.5 | 0.460 |
| Permanently sick or disabled | 2.07 (0.99, 4.31) | 0.78 | 0.053 |
| Retired | 1.41 (0.86, 2.32) | 0.36 | 0.171 |
| Looking after home or family | 0.5 (0.17, 1.44) | 0.27 | 0.199 |
| In education | - | - | - |
| Uncategorised | 1.45 (0.67, 3.17) | 0.58 | 0.348 |

NS-SEC = National Statistics Socio-economic Class.

**Table S15.** Ordinal logistic regression results for the National Child Development Study with change in stress as the outcome.

| **Variable** | **OR (95% CI)** | **SE** | ***p*** |
|---|---|---|---|
| Sex | | | |
| Male | Ref. | | |
| Female | **2.03 (1.66, 2.48)** | **0.21** | **< 0.001** |
| Ethnicity | | | |
| White | Ref. | | |
| Non-white | 0.88 (0.5, 1.56) | 0.26 | 0.662 |
| NS-SEC analytical classes | | | |
| Higher managerial | Ref. | | |
| Lower managerial | 1.01 (0.67, 1.51) | 0.21 | 0.968 |
| Intermediate occupations | 1.3 (0.9, 1.87) | 0.24 | 0.166 |
| Small employers and self employed | 0.96 (0.56, 1.64) | 0.26 | 0.887 |
| Lower supervisory and technical | 1.58 (0.93, 2.67) | 0.43 | 0.092 |
| Semi-routine occupations | 1.41 (0.93, 2.14) | 0.30 | 0.110 |
| Routine occupations | 1.14 (0.67, 1.91) | 0.30 | 0.632 |
| Uncategorised | 1.26 (0.79, 1.99) | 0.30 | 0.336 |
| Economic activity | | | |
| Employed | Ref. | | |
| Self-employed | 1.32 (0.94, 1.85) | 0.23 | 0.108 |
| Unpaid/voluntary work | 1.24 (0.48, 3.21) | 0.60 | 0.661 |
| Apprenticeship | - | - | - |
| Unemployed | 1.09 (0.51, 2.33) | 0.42 | 0.818 |
| Permanently sick or disabled | 1.45 (0.79, 2.66) | 0.45 | 0.235 |
| Retired | 1.05 (0.69, 1.6) | 0.22 | 0.809 |
| Looking after home or family | 1.04 (0.58, 1.87) | 0.31 | 0.884 |
| In education | - | - | - |
| Uncategorised | 1.60 (0.76, 3.37) | 0.61 | 0.213 |

NS-SEC = National Statistics Socio-economic Class.
